# Impact of a hospital digital coordination centre on patient flow in a large tertiary centre

**DOI:** 10.64898/2026.11.02.26367112

**Authors:** Belinda Davey, Louise Shaw, Samantha Plumb, Sarah Aranha, Timothy N Fazio, Tim Spelman, Wendy Chapman, Brad Astbury, Jana Gazarek, Peter A.D. Steel, Daniel Capurro, Portia Y. Cornell

## Abstract

Hospitals globally are under increasing strain due to rising demand and constrained capacity. This interrupted-time-series study evaluates the impact of a digital coordination centre, implemented on 28 August 2023, where a multidisciplinary operational team (comprising data scientists, patient flow managers, facilities managers, patient transport and nursing staff, for example) use real-time data dashboards showing measures of emergency department (ED) crowding and inpatient utilisation to optimise patient flow in an Australian metropolitan tertiary hospital. Analysis of 195,385 emergency department arrivals admitted to acute care and 184,454 acute inpatient episodes between July 2020 and June 2025 showed substantial improvements in measures of patient flow: 33% decrease in ED length of stay, 73% increase in hospital admissions from the ED within four hours of arrival, 8% decrease in inpatient length of stay, and 5% increase in discharges completed before noon. These changes coincided with a broader programme of organisational and operational reform, of which the DCC was a central integrating component. Hence, the findings support digitally enabled coordination centres as an effective contribution to the overall strategy for improving hospital operational efficiency.

## Introduction

Health systems internationally are under increasing pressures from rising demand, workforce shortages, and constrained hospital capacity; challenges that have been exacerbated by population ageing, patient complexity, and the COVID-19 pandemic.[1, 2] Under these conditions, impaired patient flow (the movement of patients from emergency presentation through admission, inpatient care, and discharge) leads to emergency department (ED) crowding, ED boarding (where patients are held in the ED because no inpatient beds are free), prolonged length of stay, and poorer patient outcomes. [1, 3–5] countries (including England, Canada, Ireland, New Zealand and Australia) have set targets to encourage timely admission and discharge or are considering operational strategies and funding that aim to reduce ED boarding (United States).[6, 7] Nonetheless, many hospitals continue to experience persistent bottlenecks driven by delays in inpatient capacity and discharge processes.

Given limited opportunities to increase capacity, addressing these challenges by improving internal coordination among departments and services has become a critical operational strategy for hospitals. Optimal patient flow depends on coordinated action across multiple domains, including bed management, diagnostics, discharge planning, and transport services.[8] Fragmentation and poor communication among those services limit a hospital’s ability to respond dynamically and in real time to changing demand; by contrast, timely and accurate sharing of information, supported by digital systems, can enable effective coordination and adaptive decision-making. [9–12]

To address this need, digital command and coordination centres have emerged as a model to integrate data, teams, and decision-making to improve operational awareness and patient flow. Such centres are typically centralised operational hubs in which co-located, multidisciplinary teams use real-time data from hospital systems including electronic medical records in order to provide a whole-ofhospital view of capacity and demand. Structured huddles, escalation protocols, and decision-making authority to convert shared situational awareness into coordinated action across the hospital enable teams to proactively manage patient flow,[13] often within a wider program of organisational reform. By co-locating operational functions such as bed management and patient-flow coordination with shared dashboards and interdisciplinary teams, coordination centres aim to improve situational awareness, reduce fragmentation, and accelerate coordinated responses to environmental pressures.

Early evidence suggests that coordination centres can improve operational efficiency and patient flow, including reductions in length of stay and improvements in discharge coordination. [13–16] Importantly, these centres support a shift from reactive to proactive system management by enabling real-time monitoring and predictive planning of demand and capacity.[18] However, there have been few attempts to quantify the contribution of these centres on operational efficiency. Given the investment required for these interventions, robust evaluations are needed to determine their effectiveness and inform future decision-making by policy makers and hospital management teams. The present study addresses this gap by evaluating the implementation of a Digital Coordination Centre (DCC) at the Royal Melbourne Hospital (RMH). We conducted an interrupted-timeseries study to estimate changes in key ED, inpatient, and escalation metrics after DCC implementation. We used ‘seasonal autoregressive integrated moving average time-series analyses with an exogenous variable’ (SARIMAX) estimators to adjust for pre-existing trends, seasonality and autocorrelation.

## Results

### Descriptive statistics

Table 1 shows the descriptive statistics for each of the metrics both pre and post DCC implementation. The descriptive statistics are consistent with improvements in all metrics other than the percentage of 15-minute blocks where the Ambulance Victoria or wait-room queue was between 6 and 19, or greater than 19, as this appears to have increased post implementation rather than reduced. However, the descriptive statistics should not be interpreted independently of the slopes presented in the inferential analyses below.

**Table 1:** Patient flow metrics before and after implementation.

| Patient flow metric | Pre-implementation |  | Post-implementation |  |
|---|---|---|---|---|
|  | Mean (SD) | Range | Mean (SD) | Range |
| Ambulance Vic & waiting room queue | *n* = 96,879 15-min blocks<br>*N* = 156 weeks |  | *n* = 46,266 15-min blocks<br>*N* = 79 weeks |  |
| 6–19 | 30.64 (13.60) | 0.30, 66.92 | 35.79 (12.79) | 4.17, 64.96 |
| 20+ | 1.10 (1.84) | 0.00, 8.18 | 3.13 (3.65) | 0.00, 17.11 |
| Bed variance | *n* = 104,339 15-min blocks<br>*N* = 157 weeks |  | *n* = 46,266 15-min blocks<br>*N* = 79 weeks |  |
| 10 to 19 | 17.40 (9.12) | 2.98, 43.75 | 25.54 (10.12) | 7.01, 56.55 |
| 20+ | 62.36 (16.56) | 25.74, 90.48 | 37.22 (13.24) | 8.98, 74.55 |
| Emergency department outcomes | *n* = 126,094 presentations<br>*N* = 164 weeks |  | *n* = 69,291 presentations<br>*N* = 79 weeks |  |
| Weekly mean ED length of stay (hours) | 5.92 (0.99) | 4.04, 9.40 | 5.26 (0.50) | 4.22, 6.34 |
| Weekly % admitted within 4 hours | 43.58 (7.48) | 29.95, 65.95 | 47.24 (4.32) | 39.05, 58.58 |
| Inpatient outcomes |  |  |  |  |
| Weekly mean inpatient length of stay (days) | *n* = 119,373 presentations<br>*N* = 165 weeks |  | *n* = 65,081 presentations<br>*N* = 78 weeks |  |
|  | 4.21 (0.24) | 3.51, 4.93 | 3.90 (0.28) | 2.65, 4.65 |
| Weekly % discharged by 12pm | *n* = 131,817 presentations<br>*N* = 164 weeks |  | *n* = 72,221 presentations<br>*N* = 79 weeks |  |
|  | 26.65 (2.48) | 17.69, 31.90 | 29.37 (2.19) | 20.55, 32.84 |

*Notes:* ED – emergency department; SD – standard deviation.

### Results of SARIMAX analysis

Table 2 shows the overall results of the adjusted analyses for all metrics. The overall trends are displayed in figures, and interpreted for each metric individually, following the table. Results of the sensitivity analyses using an early start date for the DCC implementation period (commencing on 13 February 2023 instead of 28 August 2023) are provided in Supplementary materials (S4).

**Table 2:** Results.

|  | **Average weekly change (95% CI)** |  | **Intervention effect (95% CI)** |  |  |  |  |
|---|---|---|---|---|---|---|---|
|  | **Pre-DCC** | **Post-DCC** | **Difference in slopes (CI)** | **Pr\|*H*<sub>0</sub>** | **Level change** | **Pr\|*H*<sub>0</sub>** | **Percent change (annualised difference)** |
| **Escalation metrics** |  |  |  |  |  |  |  |
| Ambulance Vic & waiting room queue of 6–19 | 0.17 (0.14, 0.21) | −0.10 (−0.19, −0.006) | −0.27 (−0.37, −0.17) | <0.001 | −5.07 (−10.50, 0.35) | 0.067 | 30% decrease (∼5,400 fewer triggers/year) |
| Ambulance Vic & waiting room queue of 20+ | 0.01 (0.01, 0.02) | 0.01 (−0.02, 0.03) | −0.01 (−0.03, 0.01) | 0.499 | 0.68 (−0.46, 1.82) | 0.242 | 15% increase (∼140 additional triggers/year) |
| Bed variance 10–19 | 0.13 (0.12, 0.14) | −0.10 (−0.14, −0.06) | −0.23 (−0.27, −0.19) | <0.001 | 1.07 (−1.05, 3.19) | 0.320 | 2% decrease (∼2,700 fewer triggers/year) |
| Bed variance 20+ | −0.26 (−0.28, −0.24) | −0.23 (−0.28, −0.18) | 0.03 (−0.02, 0.09) | 0.239 | 6.23 (3.06, 9.41) | <0.001 | 25% increase (∼2,600 additional triggers/year) |
| **Emergency department metrics** |  |  |  |  |  |  |  |
| Weekly mean ED length of stay (hours) | 0.01 (0.01, 0.02) | −0.005 (−0.007, −0.001) | −0.02 (−0.02, −0.02) | <0.001 | −1.84 (−2.10, −1.58) | <0.001 | 33% decrease (∼119,000 fewer hours/year) |
| Weekly % admitted within 4 hours | −0.12 (−0.13, −0.11) | 0.006 (−0.007, 0.032) | 0.12 (0.10, 0.14) | <0.001 | 15.02 (13.52, 16.52) | <0.001 | 73% increase (∼9,100 additional timely admissions/year) |
| **Inpatient metrics** |  |  |  |  |  |  |  |
| Weekly mean inpatient length of stay (days) | 0.0001 (−0.0003, 0.0005) | −0.0042 (−0.0054, −0.0030) | −0.0043 (−0.0055, −0.0030) | <0.001 | −0.16 (−0.22, −0.09) | <0.001 | 8% decrease (∼13,900 fewer inpatient days/year) |
| Weekly % discharged by 12pm | 0.011 (0.007, 0.016) | 0.02 (0.01, 0.03) | 0.01 (0.00, 0.02) | 0.018 | 0.90 (0.28, 1.51) | 0.004 | 5% increase (∼700 additional early discharges/year) |

*Notes:* ED – emergency department; NS – not significant; Post-DCC – Post-implementation period; CI – 95% confidence interval; Pr\|*H*<sub>0</sub> – probability of the observed or more extreme results, given that the null hypothesis is true.

### Ambulance Victoria and waiting room queue

The weekly average percent of 15-minute blocks where the combined Ambulance Victoria and ED waiting room queues were 6 to 19 patients is plotted in Figure 1. The positive increasing rate of 0.17 percentage points per week (CI: 0.14 to 0.21), on average, prior to DCC implementation was attenuated to −0.10 percentage points per week (CI: −0.19 to −0.006), on average, after DCC implementation, resulting in a significant average difference of −0.27 percentage points (p<0.001, 95% CI: −0.37 to −0.17). The level change was not significant (p=0.067).

**Figure 1:**
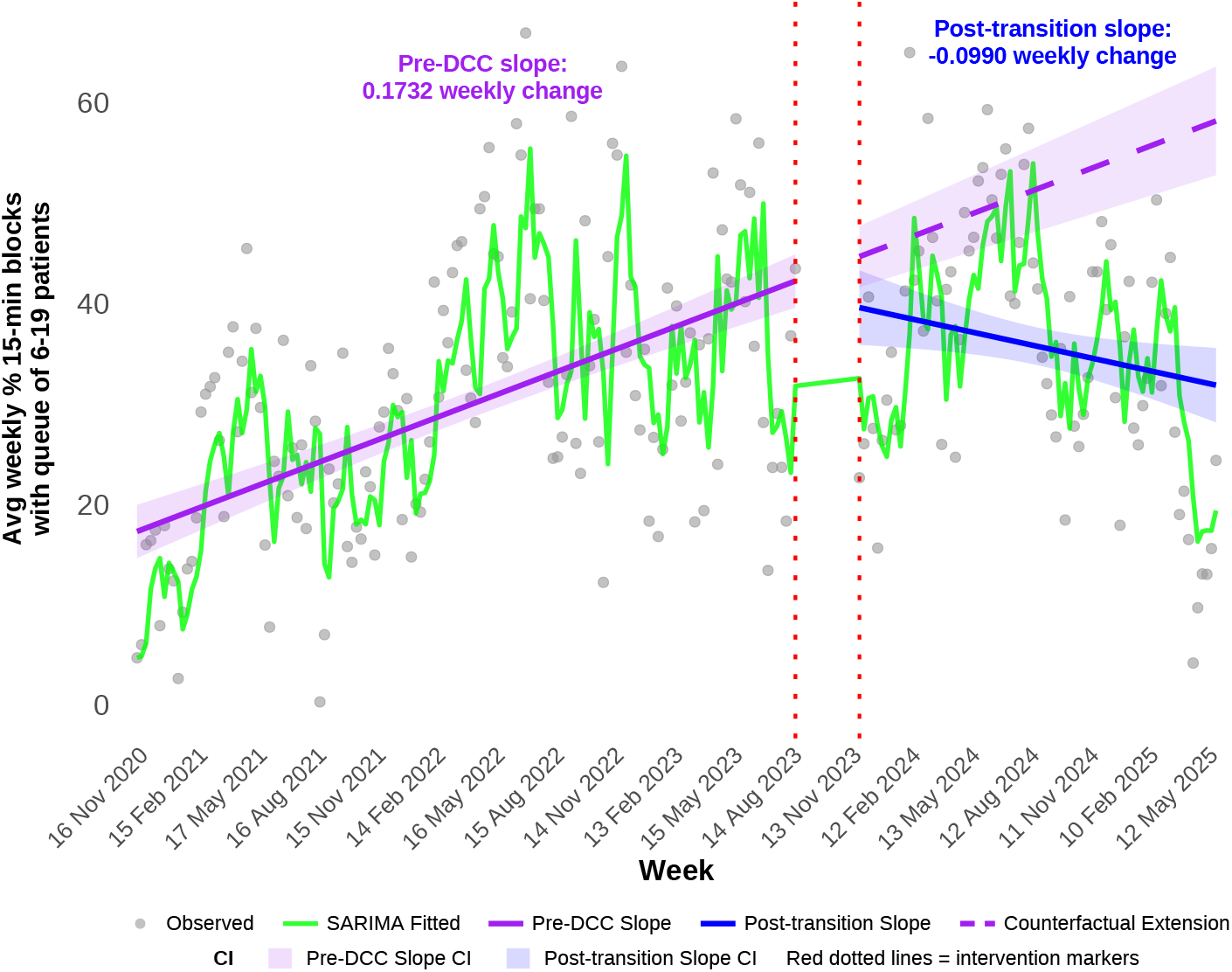
Change in weekly percent of 15-minute blocks where the Ambulance Victoria and waiting room queues reached 6 to 19 patients, before and after DCC implementation; and expected values had the DCC not been implemented (the counterfactual trend)

The DCC improvements correspond with approximately 5,400 fewer 15-minute blocks per year on average that reached the escalation trigger of 6-19 across the post-implementation period, or ∼100 fewer per week, or about 20 fewer per day i.e., a 30% reduction.

Figure 2 displays the weekly percentage of 15-minute blocks where the total Ambulance Victoria and waiting room queue was 20 or greater. The slope and level changes were not statistically significant.

**Figure 2:**
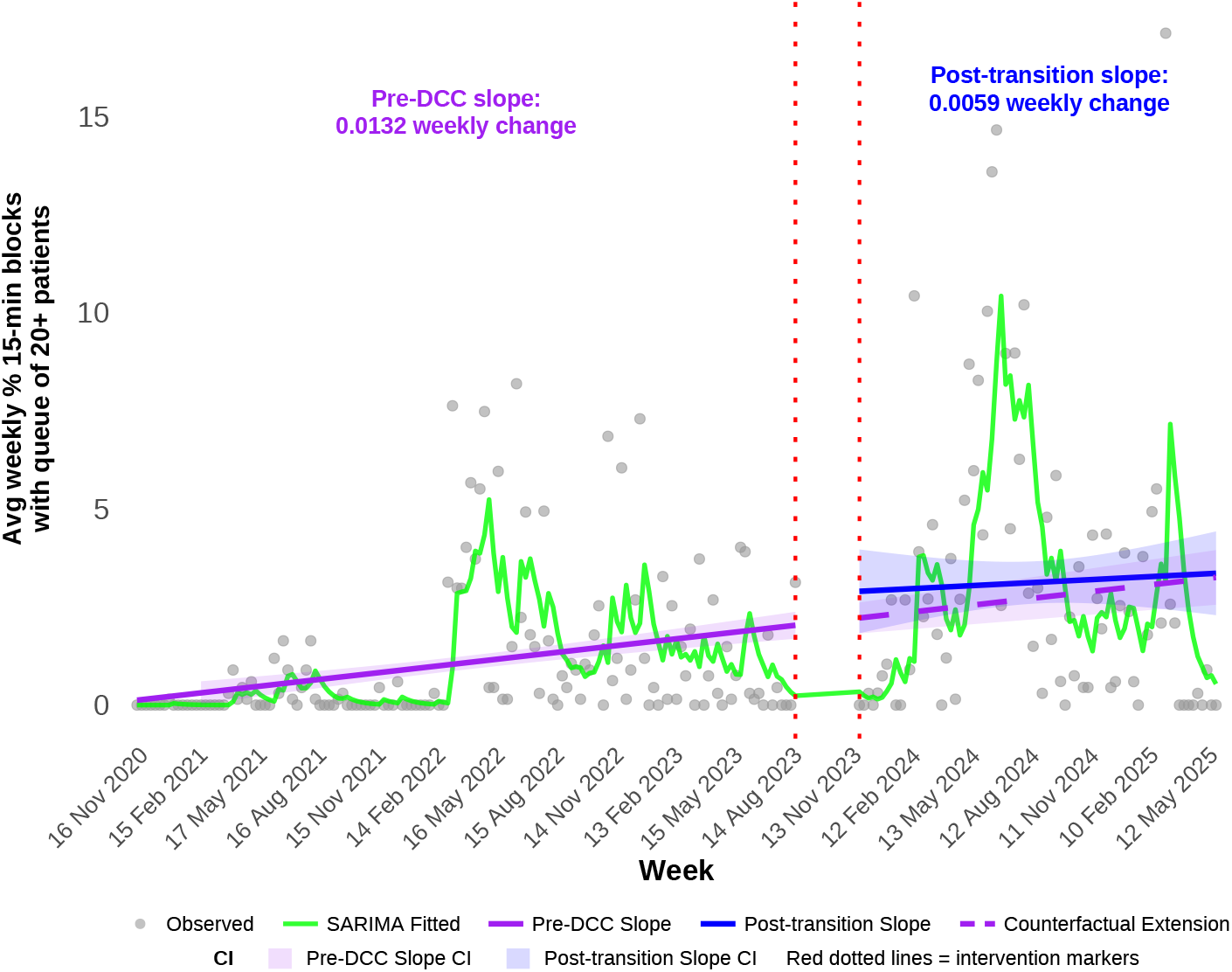
Change in weekly percent of 15-minute blocks where the Ambulance Victoria and waiting room queues reached 20 or more patients, before and after DCC implementation; and expected values had the DCC not been implemented (the counterfactual trend)

### Bed variance

The average weekly percentage of 15-minute blocks where bed variance was in the 10 to 19 range are plotted in Figure 3. The increasing rate of 0.14 percentage points per week (CI: 0.12, 0.14), on average, prior to DCC implementation was converted to a decreasing rate of 0.10 percentage points per week (CI: −0.14, −0.06), on average, after DCC implementation. This is a significant change in weekly trend of 0.23 percentage points per week (CI: −0.27, −0.19, p<.001). No statistically significant change in the level immediately before and after the implementation was detected.

**Figure 3:**
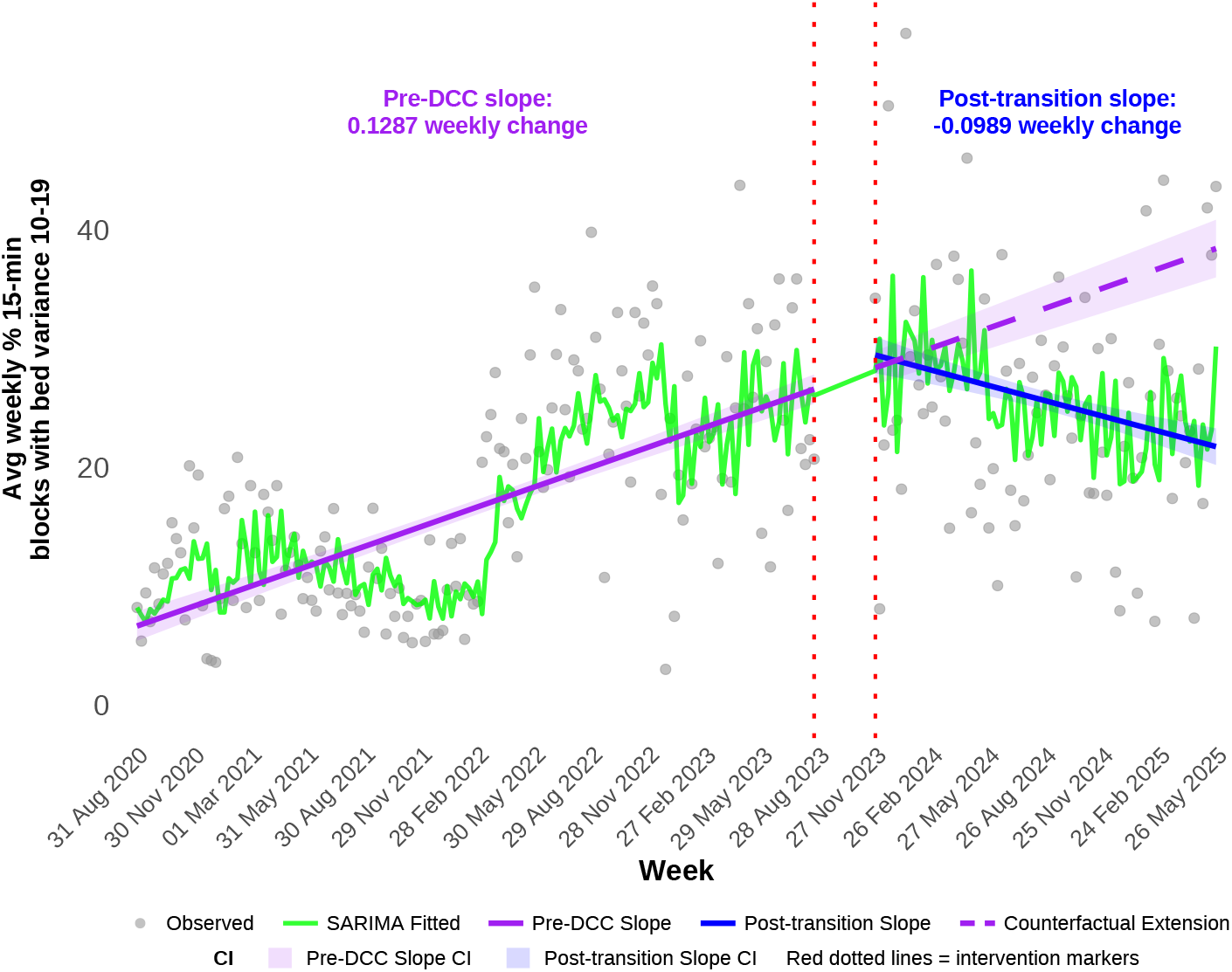
Change in the weekly mean percentage of 15-minute blocks where bed variance was 10 to 19 patients, before and after DCC implementation, and expected values had the DCC not been implemented (the counterfactual trend)

The DCC improvements correspond with 2,700 fewer 15-minute blocks that reached this escalation trigger annually, a 24% reduction compared to the forecast.

In regard to the percent of 15-minute blocks with bed variance of 20 or more, there was no evidence of a statistically significant difference in the pre-DCC and postimplementation trends (p=.239); (see Figure 4). However, there was a significant increase in the level change from the pre-DCC to the post-implementation phase of 6.23 percentage points (95%CI: 3.06, 9,41; p<0.001). As there are 672 blocks per week, this level change corresponds to an annual increase of ∼2,600 additional escalation triggers per year in this range, a 25% difference.

**Figure 4:**
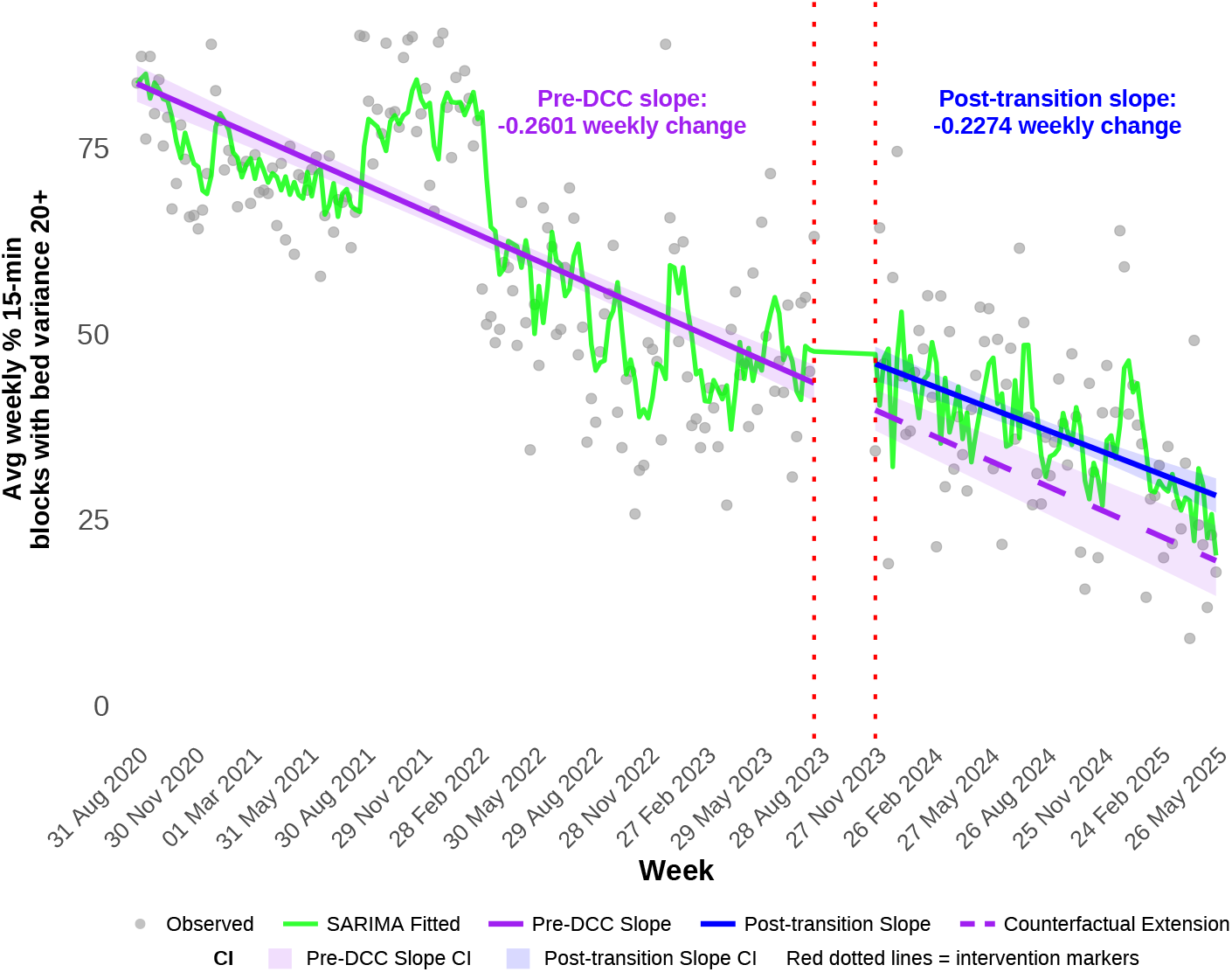
Change in weekly percentage for 15-minute blocks where bed variance was 20 or greater, before and after DCC implementation, and expected values had the DCC not been implemented (the counterfactual trend)

### Length of stay for admitted ED patients

The average weekly mean ED length of stay (hours) is plotted in Figure 5. For admitted ED patients, the mean ED length of stay (LoS) was increasing by 0.015 hours each week prior to DCC implementation (CI: 0.013, 0.016). That trend reversed after DCC implementation to a decreasing trend of 0.005 hours per week (CI: −0.007, −0.002). There was also a significant level change of -1.84 hours per patient (p<0.001, CI: -2.10, -1.58), from the end of the pre-DCC period to the beginning of the post-implementation period. The DCC improvements correspond with a saving of approximately 119,000 hours (∼5,000 days) spent waiting in ED by patients annually, a 33% improvement.

**Figure 5:**
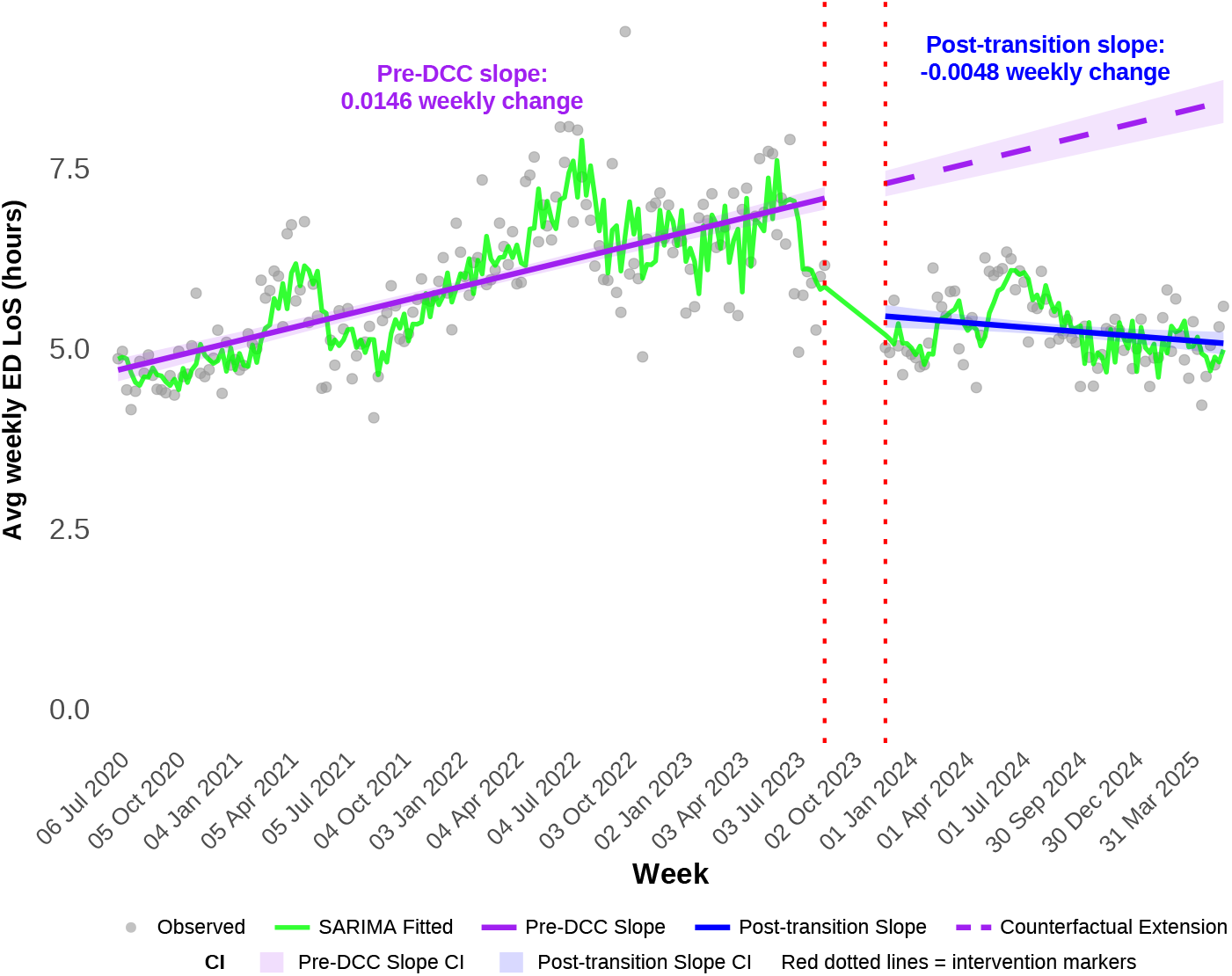
Change in weekly average emergency department length of stay per week, before and after DCC implementation, and expected values had the DCC not been implemented (the counterfactual trend)

### Percent of admissions within 4 hours

Figure 6 shows the average weekly percent of admissions within 4 hours from arrival at the ED. This measure decreased at a rate of 0.12 percentage points per week (CI: −0.13, −0.11) prior to DCC implementation, which was attenuated after DCC implementation to 0.006 (CI: −0.017, 0.018), a significant improvement of 0.12 percentage points per week (p<0.001, CI: 0.10, 0.14). There was also an immediate increase from the end of the pre-DCC period to the beginning of the post-implementation period of 15.02 percentage points (CI: 13.52, 16.52, p<.001).

**Figure 6:**
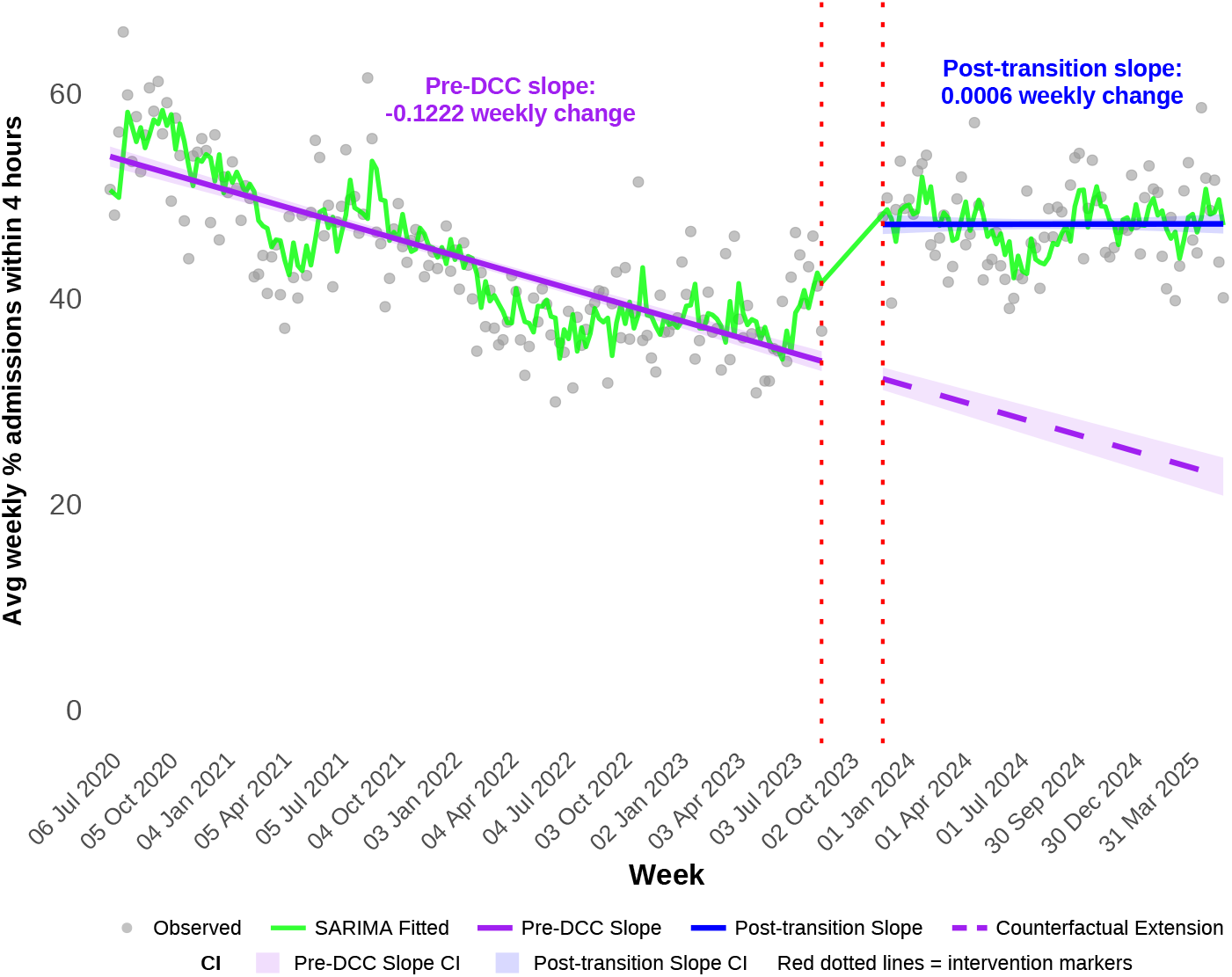
Change in weekly percentage of ED patients admitted within 4 hours of presentation, before and after DCC implementation, and expected values had the DCC not been implemented (the counterfactual trend)

The DCC improvements correspond with approximately 9,100 additional timely admissions from ED per year (∼170 per week), a 73% increase.

### Average weekly inpatient length of stay

The trend in inpatient length of stay changed from a flat rate of 0.0001 days per episode per week (CI: −0.0003 days, 0.0005 days) to decreasing at a rate of 0.004 days per episode per week (CI: −0.005, −0.003), a difference in the rate of change of 0.004 fewer days per episode each week (CI: −0.006, −0.003, p<.001). The level change using the linear model also differed significantly from the end of the pre-DCC period to the start of the post-implementation period, with an immediate reduction of 0.16 days per episode (95% CI: −0.22, −0.09; p<0.001). This is displayed in Figure 7.

**Figure 7:**
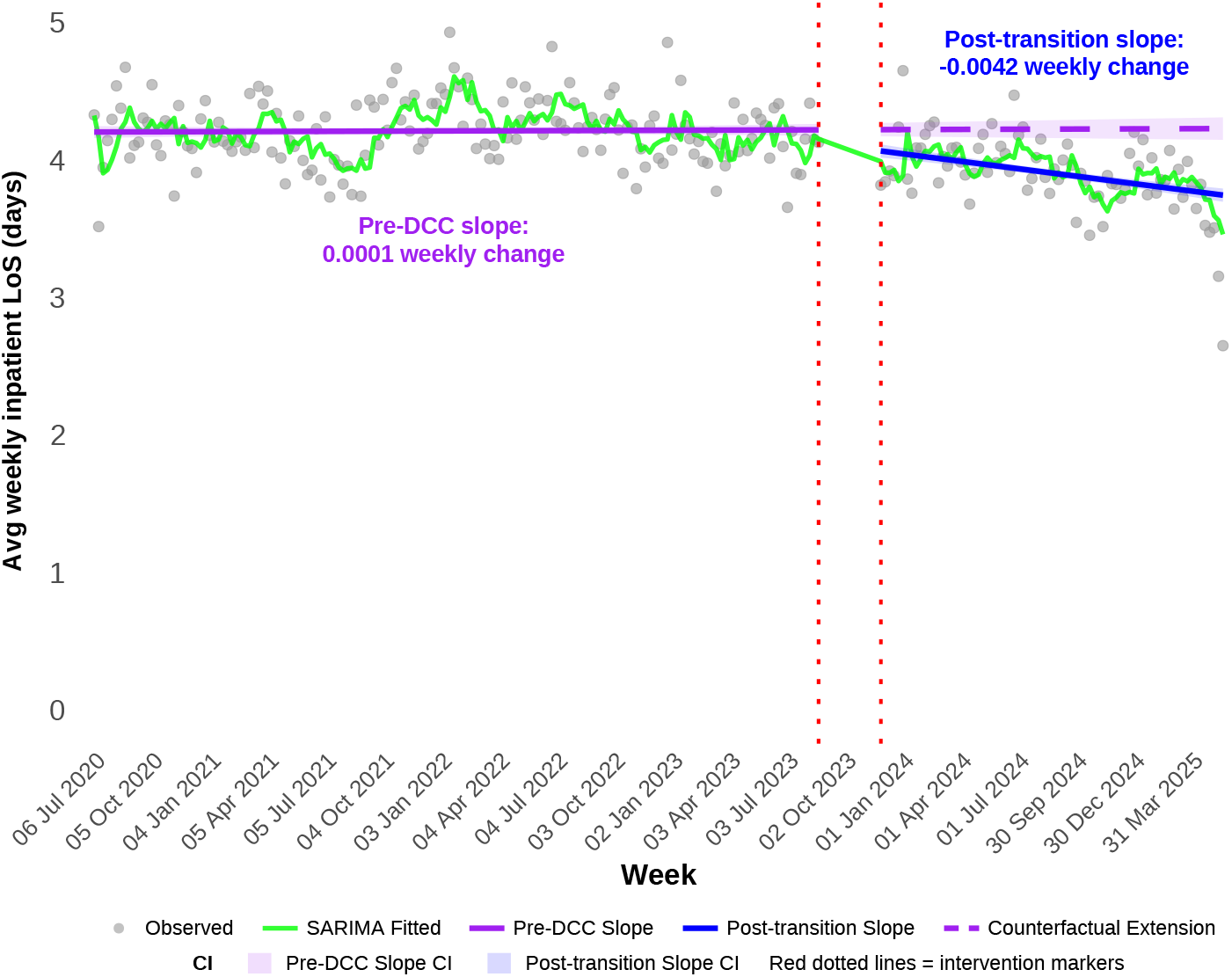
Change over time in average weekly inpatient length of stay before and after DCC implementation, and expected values had the DCC not been implemented (counterfactual trend)

The decrease in length of stay equates to approximately 13,800 fewer acute inpatient days per year, an 8% improvement compared to the prediction without the DCC.

### Percent of discharges by 12pm

The trend in percentage of timely discharges (prior to 12pm) was increasing slightly prior to the DCC implementation at a rate of 0.011 percentage points per week (CI: 0.007, 0.016) and increased at a higher rate to 0.02 percentage points per week (CI: 0.01, 0.03) post-DCC implementation, a difference in the rate of increase of 0.01 percentage points per week (CI: 0.002, 0.024, p=0.018; see Figure 8). The level change using the linear model also differed significantly from the end of the pre-DCC period to the start of the post-implementation period, with an immediate increase of 0.90 percentage points in discharges prior to 12pm (95% CI: 0.28, 1.51; p=0.004). The change in rate suggests there were 690 additional timely discharges by 12pm per year in the post-implementation period, a 5% improvement compared to the projected number.

**Figure 8:**
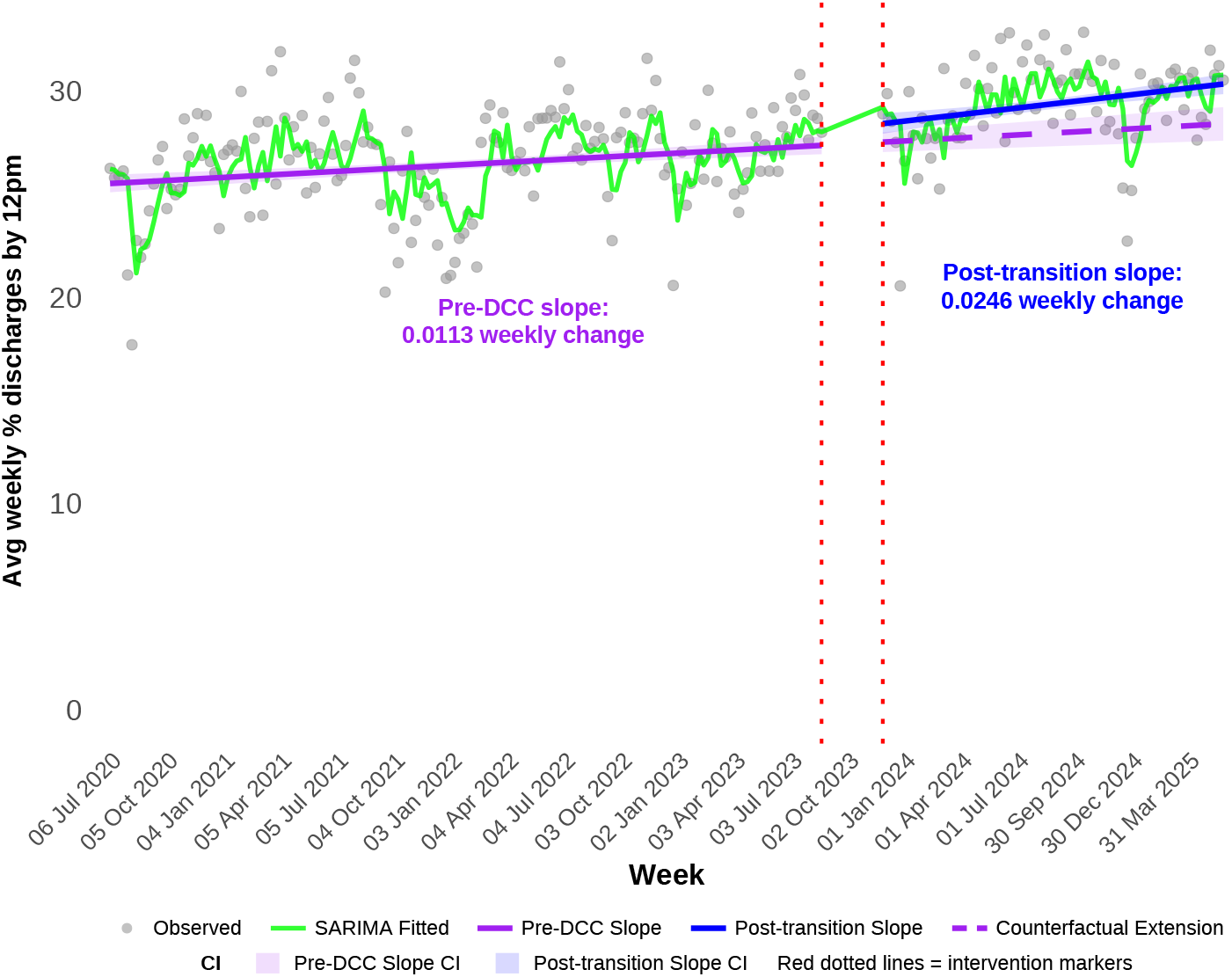
Change over time in average weekly inpatient length of stay before and after DCC implementation, and expected values had the DCC not been implemented (counterfactual trend)

## Discussion

This study evaluated the contribution of implementing a Digital Coordination Centre (DCC) at a large tertiary hospital using interrupted-time-series methods. DCC implementation was associated with significant operational improvements in several patient-flow outcomes, including reduced ED length of stay for admitted patients, increased proportion of ED admissions occurring within four hours, reduced acute inpatient length of stay, and increased discharges before noon. When translated into operational units, these changes corresponded to substantial annualised capacity gains: approximately 120,000 fewer ED hours, 9,100 additional timely ED admissions, 14,000 fewer acute inpatient bed-days, and 700 additional discharges before noon per year. As noted in the design and setting, the DCC was implemented alongside, and helped operationalise, a broader package of organisational changes, including a centralised Patient Flow Coordinator team, a purpose-built transit lounge with real-time digital workflows, and general medicine bed and model-of-care reforms. Hence, the DCC represents an integrating mechanism for these changes, rather than standing as an isolated intervention. Indeed, the DCC may function as a visible manifestation of a broader shift in organisational priorities rather than the sole causal mechanism.

The escalation metrics provide a more nuanced interpretation. Moderate operational-strain thresholds improved for both bed variance and the combined Ambulance Victoria / waiting room queue. In contrast, the most severe thresholds did not consistently improve. This pattern of improvement at moderate thresholds, and no improvement at the most severe threshold, is consistent with a coordination centre optimising the use of latent capacity rather than creating new capacity, for example, accelerating bed allocation, prioritising discharges, coordinating cross-functional action. At extreme demand, when structural constraints dominate, real-time coordination is unlikely to be sufficient. These findings suggest that DCCs should be understood as capacity-optimising interventions, not substitutes for adequate beds, staffing, and downstream care capacity.

A slight level increase in the higher bed variance threshold was observed after DCC implementation, though a declining trend continued. The slight increase may represent some reversion to the mean, or it may be due to improved recording fidelity after DCC implementation. That is, the DCC itself may introduce measurement bias by improving recording fidelity, especially given the improved tracking of escalation metrics in response to DCC implementation. Improved recording fidelity is itself an operational improvement (e.g., a bed marked available sooner enables earlier cleaning and admission).

The RMH Digital Coordination Centre was conceived and implemented in the context of burgeoning strain on hospital systems in Australia and internationally, with record high urgent presentations and ED boarding. In this context, the observed improvements are operationally and clinically meaningful because ED crowding and prolonged boarding are associated with delayed care, worse outcomes, and increased length of stay.[19–21]

Previous studies have reported improvements in operational metrics such as ED throughput, bed utilisation, and discharge coordination following implementation of similar models. However, previous literature has relied on descriptive analyses,[9, 22, 23] has not taken the time-series structure or seasonality into account,[14] have included data during the COVID-19 pandemic, [11, 15] or have focused on the ICU,[14]—limiting their ability to draw conclusions about the hospital-wide contribution of a command-centre intervention. Moreover, differences in hospital organisation and processes within and between countries makes comparison of outcomes difficult across studies.[24]

The results align with broader systems-engineering perspectives on healthcare operations.[9] Hospitals function as complex sociotechnical systems in which delays or inefficiencies in one area can propagate throughout the system. Command or coordination centres attempt to address this complexity by integrating operational data, facilitating multidisciplinary collaboration, and enabling proactive management of system pressures.[18] The results from this study support the hypothesis that digitally enabled coordination may contribute to improvements in hospital flow when dashboards, co-located expertise, escalation protocols, and executive decision-making are integrated into daily operations.

The present findings are broadly consistent with reports from previous hospital command-centre evaluations, which have described improvements in measures including bed allocation, transfers, emergency department boarding, and other aspects of patient flow. A scoping review of hospital capacity command centres found that all included studies showed improvement in at least some performance measures, although considerable heterogeneity in command-centre design and evidence quality limits the generalisability of these findings.[25] For example, implementation of the Johns Hopkins Capacity Command Center was associated with reduced patient-flow delays despite an increase in hospital occupancy.[9] However, not all evaluations have demonstrated consistent improvements. An interrupted time-series evaluation at Bradford Royal Infirmary in the UK found modest improvements in some stages of emergency department flow but no significant or consistent improvement in overall length of stay or several other flow measures.[11] Differences in command-centre functions, baseline digital maturity, concurrent improvement initiatives, healthcare-system context, outcome definitions, and study designs substantially limit direct comparison between reported effect sizes. The findings of the present study should therefore be interpreted within their particular organisational and health-system context rather than assumed to generalise to all command-centre implementations.

This study has several strengths that support the credibility of its findings. First, the analysis used a long observation period spanning more than three years prior to implementation and over one year following the establishment of the DCC. This extended time frame allowed underlying temporal trends to be identified and statistically adjusted in the modelling process, making it possible to compare observed post-implementation outcomes with counterfactual projections.

Second, interrupted time series analysis is a strong study design within the limitations of this kind of observational data. In the presence of steep pre-intervention trends, a crude comparison of means before and after an intervention can produce a paradoxical result where the difference in means suggests a change in one direction but the shift in level and trend suggest the opposite, as with the AV waiting room queues and bed variance escalation triggers, underlining the importance of modelling the trends over time. Furthermore, the study applied SARIMAX time-series models to account for seasonality, autocorrelation, and other statistical properties commonly present in health system data. Failing to account for these temporal features can lead to biased statistical inferences, including underestimated or inflated standard errors, incorrect confidence intervals, and inaccurate p-values, thereby increasing the risk of false-positive or false-negative findings. [26, 27] In addition, unmodelled seasonality or serial dependence may result in intervention effects being incorrectly attributed to the intervention itself when they instead reflect underlying temporal patterns in the data. [28, 29] Diagnostic testing indicated that the models adequately captured the temporal structure of the data, supporting the validity of the estimated effects. This use of time-series evaluation aligns with best-practice recommendations when randomisation is not feasible for hospital-wide interventions and data on a non-intervention comparison group are not available.[27]

Third, the evaluation examined multiple complementary outcomes across both ED and inpatient domains. Improvements observed across several related metrics strengthen confidence that the intervention affected system-level patient flow rather than producing isolated or artefactual changes in individual measures. The inclusion of escalation-trigger analyses is a particular point of novelty because they link outcomes to operational “strain” indicators that plausibly mediate patient flow and are conceptually consistent with the real-time threshold logic embedded in DCC or command-centre interventions.

Finally, the analysis translated statistical findings into operationally meaningful estimates, such as total hours saved in the ED and total inpatient days avoided. These estimates are important for executive decision-making, because they connect statistical findings to capacity, service delivery, and potential return on investment.

Several limitations should also be considered when interpreting these findings. First, it cannot be definitively concluded that the observed improvements are solely attributable to the DCC. Other concurrent organisational initiatives, changes in staffing, or broader health-system trends may also have contributed to the observed changes. This is partly a limitation of conducting a single-site study. As noted in design and setting, multiple organisational initiatives focused on improving inpatient access were implemented during the study period. These initiatives may also account for the improvements in metrics observed throughout the study. Nonetheless, the DCC functions as an integrating hub that aligns, coordinates and amplifies the impact of these initiatives, making it a critical enabling mechanism underpinning system-wide improvements in patient access and flow.

Second, though our results were generally robust to different modelling choices in overall direction, some choices inevitably affect the size of the estimates. While the study excluded the initial months of the COVID-19 pandemic with the most drastic differences in activity observed in the data, Victoria continued to experience restrictions and altered patterns of hospital use into 2021. To the degree that restrictions suppressed emergency department presentations and hospitalisations, inclusion of this period may have exaggerated pre-implementation trends, though the volatility of the ED and inpatient metrics measured in this study had reduced substantially by 6 July 2020, when data analysis began for this study (see the average weekly ED and inpatient arrivals from 2020 to 2026 in the Supplementary materials, S3). The annualised percentage-change estimates are calculated under the assumption that the counterfactual is a continuation of the pre-implementation trend. While modelling the counterfactual on observed data is a defensible assumption, assuming a flat trend for the counterfactual would have produced more conservative percentage-change estimates.

Second, the analysis focused on operational performance metrics and did not examine patient-centred outcomes such as patient experience, safety outcomes, or clinical quality indicators. Although improved patient flow is expected to have downstream benefits for patients and providers [3–5], further research is needed to confirm these effects. Moreover, while reducing unnecessary time in hospital is beneficial, hospitals must take care not to reduce necessary stays. Inclusion of patient-centred outcomes would examine this.

Third, some operational metrics, particularly escalation indicators, may be influenced by factors external to hospital control, such as community demand, ambulance availability, or seasonal variations in illness patterns. Although the time-series models adjusted for temporal patterns, these external factors remain important contextual considerations; command-centre evaluations are conducted within complex and dynamic settings where attribution must be interpreted with appropriate caution.

Finally, the observed effects likely reflect the combined influence of data infrastructure, dashboard design, co-located operational teams, governance routines, escalation protocols, and cultural change. This study was not designed to isolate the marginal contribution of each component. Moreover, the particular design and implementation of coordination centres within hospitals are necessarily specific to their organisational context and operational setting. Hence, providing evidence that this particular implementation was successful may not generalise to the implementation of coordination centres elsewhere.

Despite these limitations, the findings have important implications for hospital management and health system policy. Digital coordination centres may improve patient flow by integrating real-time data with multidisciplinary operational teams, supporting a shift from reactive crisis management toward proactive system coordination.[18] However, technological infrastructure alone is unlikely to be sufficient. Improvements depend on embedding dashboards within governance structures, escalation processes, clearly defined responsibilities, and models of care that translate information into timely, coordinated action.[30] Digital coordination centres should therefore be understood as sociotechnical and organisational interventions rather than simply digital dashboards.

Future research should examine whether these findings are replicable across different hospital settings using stronger designs, including controlled interrupted time series, stepped-wedge approaches, or, where feasible, site-level randomisation. Evaluations should also incorporate measures of case mix, acuity, comorbidity, diagnostic complexity, and service demand to better account for time-varying confounding and determine whether improvements persist across differing levels of system pressure[31, 32]. This is particularly important given the mixed command-centre evidence base and heterogeneity in implementation and study design.[25]

Further work should investigate the mechanisms through which coordination centres affect hospital operations, including communication, decision-making, workflows, and organisational culture. Mixed-methods studies involving staff and patients could support development of a theoretical framework linking DCC processes to operational and patient outcomes [25, 33]. Evaluations should include patient-centred measures such as re-presentations, readmissions, adverse events, mortality, and PROMs/PREMs to ensure improvements in flow do not occur at the expense of care quality.[34, 35]

Predictive analytics may provide an additional opportunity to strengthen DCC function. Models using historical and real-time hospital data can forecast demand, admission risk, prolonged length of stay, and bed availability, potentially enabling earlier intervention before bottlenecks develop.[36–38, 41**? ?** ] However, their value will similarly depend on governance and workflows that translate predictions into operational action.

Finally, economic evaluation is needed to determine the DCC’s return on investment and broader health-system value. Future studies should estimate savings associated with reduced length of stay, improved bed turnover, and reduced ED boarding, alongside cost-utility analyses incorporating quality-adjusted life years where appropriate. Such evidence would assist health-system leaders and policymakers in determining whether digital coordination centres warrant wider implementation.

In summary, the implementation of a Digital Coordination Centre at a large tertiary hospital was associated with significant improvements in several key patient-flow metrics, including emergency department throughput, inpatient length of stay, and discharge timing. These improvements suggest that digitally enabled coordination models may contribute to more efficient hospital operations in increasingly pressured healthcare systems. While further research is needed to confirm these findings in other settings and to understand the mechanisms underlying these improvements, the results contribute valuable evidence to the emerging literature on hospital command and coordination centres.

## Methods

### Design & setting

This study is a single-site, retrospective, segmented interrupted-time-series study of the Digital Coordination Centre (DCC), a hospital-wide operational hub designed to improve patient flow, capacity management, and coordination of care across the Royal Melbourne Hospital (RMH), a public, tertiary referral teaching hospital in Victoria, Australia. The RMH is a large public tertiary/quaternary referral and teaching hospital providing both general and highly specialised services, including major trauma, cardiac, neuroscience, oncology, intensive care, and infectious diseases services. It receives referrals from metropolitan, regional and rural Victoria, as well as interstate, managing a complex case mix and substantial demand for specialist and acute inpatient capacity. The hospital operates within Victoria’s publicly funded health system, in which hospital activity and capacity must be managed within government funding and resource allocations. This context differs from predominantly private or insurance-funded hospital systems, such as that of the United States, and places considerable emphasis on optimising the use of finite public hospital beds, workforce, and other resources.

The DCC integrates real-time data from electronic medical records and hospital information systems into digital dashboards that provide a whole-of-hospital view of operational status, including emergency department activity, inpatient bed availability, predicted discharges, escalation triggers, and patient-flow pressures. This digital context is also relevant to the generalisability of the intervention. Adoption and maturity of electronic medical records and interoperable clinical information systems remain variable across Australian hospitals, meaning that not all health services have equivalent access to the integrated, near-realtime data infrastructure required to reproduce the RMH model. Consequently, implementation of a comparable coordination centre elsewhere may require substantial preceding investment in digital infrastructure, data integration, and data governance.

Importantly, the DCC is not solely a technological intervention. Operational teams responsible for functions such as bed management, patient-flow coordination, nursing allocation, environmental services, facilities management, and transport coordination are co-located within the centre. Bringing these functions together provides a common operational picture and facilitates rapid communication and collaborative decision-making across services that may otherwise operate separately. The DCC was implemented as part of a broader transformation strategy intended to shift hospital operations from reactive crisis management toward proactive, system-wide coordination. By combining integrated digital dashboards with multidisciplinary operational governance, the DCC aims to improve situational awareness, identify bottlenecks earlier, and facilitate timely interventions to optimise patient movement through the hospital. The model draws on command-centre approaches used in industries such as aviation, logistics, and emergency management, adapting these principles to the healthcare setting to support more efficient allocation of beds, staffing, and other operational resources.[42]

As this was a real-world study conducted in a non-laboratory setting, several initiatives were implemented during the study period, and some in preparation for it. For example, a Transit Lounge model-of-care was launched early in 2023 (about six months prior to DCC implementation in August 2023) that includes digital workflows and real-time dashboards, which were subsequently actively monitored by the DCC. Around the same time, a Patient Flow Co-ordinator team commenced, which subsequently operated within the DCC. As these initiatives commenced during the pre-DCC period, against which the post-DCC period is compared, the contribution of DCC implementation to patient outcomes may be attenuated. Hence, this intervention could not be isolated from several other initiatives, some of which were implemented during the study period and some in preparation for it.

On the other hand, the ‘fast track and rapid stay’ model of care, operationalised in the ED in January 2024, was focused on flow of non-admitted patients and is therefore unlikely to have had a strong effect on metrics reported in this analysis, as these focus on admitted patients (see ‘Participants and eligibility criteria’ below). Similarly, the Victorian Virtual ED (an initiative external to RMH) focused on non-admitted patients, and began during the pre-DCC period, so will likely have similar impacts upon the pre-DCC and post-implementation periods.

A retrospective longitudinal time-series evaluation using ‘seasonal autoregressive integrated moving average models with exogenous intervention terms’ (SARIMAX; described further in ‘Statistical analyses’) was undertaken as part of a mixed-methods evaluation of the impact of introducing the DCC at the Royal Melbourne Hospital. The study was approved by the hospital’s human research ethics committee (protocol no. QA2023089). This paper reports the impact of the DCC introduction on patient flow metrics, constituting the quantitative component of the multi-methods evaluation.[42, 43]

### Data and observation periods

Data were extracted from the hospital’s data warehouse on 3 July 2025. Bed variance and escalation-metrics data were available from 17 August 2020. However, escalation metric values were relatively low for the first couple of weeks of data collection for bed variance and until half-way through November for the Ambulance Victoria and wait room metrics (see Figure S1 and S2 in Supplementary Material S1 to see an illustration of these values in the time series). These low values appear to be the result of operational factors where hospital staff took some time to regularly report escalation metrics. In order to ensure that the impact of the DCC intervention is not over estimated, the pre-DCC period was set to start on 31 August 2020 for bed variance and 16 November 2020 for the Ambulance Victoria and waiting room queues.

The remaining metrics were available from 2017 onwards. However, because early COVID-19 pandemic restrictions in Victoria suppressed emergency department presentations and hospital activity, only data after 6 July 2020 were used in analyses;[44] while COVID-19 lockdowns continued in Victoria beyond this time, examination of the metrics in time-series shows a reduction in marked volatility of the data during the COVID-19 period by July, 2020 (see Supplementary materials, S2). Measures were aggregated as weekly averages to address weekly fluctuation patterns and day-to-day noise while preserving sufficient temporal resolution for interrupted-time-series modelling.

Weeks were assigned to a pre-implementation period (from 6 July 2020 to 27 August 2023) and a post-implementation period (from Monday, 27 November 2023 to Monday, 30 June 2025). A 13-week transition period (from 28 August 2023 to 26 November 2023), when the DCC was being implemented and staff were being trained, was excluded from analyses.

Note, however, that as some aspects of the DCC implementation were implemented in early 2023 (specifically, a Patient Flow Co-ordinator team was introduced, a purpose-built transit lounge was opened, and digital workflows and real-time dashboards were implemented) a sensitivity analysis was conducted whereby the DCC implementation period was set to Monday, 13 February 2023, instead of 28 August 2023.

### Outcomes

Two emergency department (ED) outcomes were analysed for presentations leading to acute hospital admissions: length of stay in the ED (the time in hours a patient spent in the ED from arrival to inpatient admission) and the proportion of ED patients admitted to an inpatient ward within four hours of presentation. The latter aligns with the four-hour National Emergency Access Target (NEAT) used in Australian public hospital performance reporting.[45] For inpatient metrics, length of stay and percent of discharges completed prior to 12pm were included. Greater proportion of inpatient discharges completed before midday may increase bed availability and reduce ED boarding, though recent evidence is mixed.[10, 46]

Two metrics related to ED boarding and representing sufficient bed capacity to meet demand were examined. When the measures reach certain thresholds— informed by historical data and developed over an 18-month period through consultation with informatics and clinical operations teams prior to the DCC implementation — they create ‘escalation triggers’. These triggers prompt the operations teams to form decision-making huddles and consider actions such as opening of closed and transit lounge beds, bringing in additional staff, and prioritising (safe) discharges.

The first escalation trigger, ‘bed variance,’ is the anticipated number of required admissions calculated from historical averages, minus unallocated beds (according to a midnight census) and discharges over the course of the day. The second escalation trigger, Ambulance Victoria (AV) / waiting room queue, is the number of patients being attended to by ambulance crews in corridors plus patients in the waiting room queue waiting to be brought into an acute treatment area in the ED (further details regarding the capacity management levels associated with escalation triggers are provided in the Supplementary materials, S2). The AV/waiting room queue is a measure of ED boarding that results in a delay to definitive emergency care and can affect ambulance response times and community safety.[47]

Both metrics were recorded in 15-minute increments and displayed in the DCC dashboards as threshold indicators: for bed variance, escalation thresholds were 10 to 19 (amber) and 20 or more (red) (Hospital) deficit of beds (i.e. bed variance); and for ambulance/waiting room queues, 6 to 19 (amber) and 20 or more (red) (Hospital) patients waiting for a treatment space. These categories were selected because they correspond to operationally meaningful escalation thresholds used and displayed separately in the Digital Coordination Centre (DCC) dashboards. The analysis examined the average weekly proportion of 15-minute increments where these thresholds were exceeded. Each category was analysed as a separate outcome, rather than using cumulative thresholds ( 10 and 20) or mean bed variance, to preserve the operational distinction between moderate and more severe capacity pressures.

### Participants and eligibility criteria

#### Emergency department

Only ED presentations linked to an inpatient admission were included in the analysis. Mental-health related, rapid-stay mental health and emergency behaviouralassessment presentations and admissions were excluded (n = 217,820) from analysis of 423,642 ED presentations, as mental-health bed access and control are managed separately from general hospital patient flow, leaving 205,822. After removing 10,437 ED arrivals during the transition period, 195,385 arrivals remained across 244 weeks. Data for these arrivals were summarised to provide weekly averages, leaving 244 data points for the ED length-of-stay and dischargebefore-noon metrics (164 in the pre-DCC period and 79 in the post-transition period).

#### Inpatient measures

Inpatient length-of-stay (LoS) and the percentage of inpatients discharged prior to 12pm were examined. These were provided for each patient admission, from which weekly averages were calculated. Only acute admissions (n=542,758) were included as the DCC is less likely to impact other types of admissions (e.g., mental health, palliative, or rehabilitative care); 22,932 non-acute admissions were excluded. Same day admissions (*i.e.,* patients discharged on the same day they were admitted) were excluded (n = 339,528) as the DCC aims to target more complex presentations with barriers to discharge. Inpatient stays from the ‘Hospital in the Home’ program were also excluded (n = 5,965), as the DCC does not manage these patients.

The DCC was not expected to impact unusually long stays as barriers to discharge are usually due to external, social factors outside of the DCC’s purview, such as waiting to appoint a guardian, or for disability housing to become available. Hence, episodes exceeding 35 days (n = 1,832) were excluded. As length of stay is logged at discharge, longer stays would not yet be recorded for the final five weeks (equivalent to excluding episodes beyond 35 days), artificially lowering average LoS at this time; hence, the final five weeks were omitted from LoS calculations (n = 515 excluded). Finally, 10,464 admissions during the transition period were excluded. This left 184,454 admissions. Weekly averages were calculated from these admissions, leaving 240 data points for each metric (164 in the pre-DCC period and 75 in the post-transition period for length-of-stay, but 79 for discharge data).

### Statistical analyses

Interrupted time-series analyses were conducted to compare the level and rate of change in each metric during each period. Specifically, seasonal autoregressive integrated moving average time-series analyses with an exogenous variable (SARIMAX) were implemented. The treatment variable is an indicator representing the post-implementation period.

SARIMAX modelling was used to account for the temporal structure of the data, including autocorrelation and seasonality, with differencing incorporated where necessary to address non-stationarity. Model adequacy was assessed through residual diagnostics, including checks for remaining serial correlation. Seasonality was assessed by comparing yearly (52-week), quarterly (13-week), and non-seasonal candidate models. Seasonal specifications were retained only when seasonal autoregressive or moving-average components were present and model fit was superior, or within 2 Akaike Information Criterion (AIC) units of the best-fitting model; otherwise, non-seasonal models were selected on grounds of parsimony. Model diagnostics and assumption tests are provided in the Supplementary Materials,S4.

The regression error is modelled as a seasonal ARIMA process:

From the SARIMAX models, the average change over time (the slope) in the weekly metrics were calculated during the pre-DCC and post-implementation periods by fitting a linear model to the SARIMAX model. The segmented regression component was specified as:

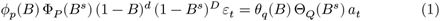

where *I_t_* identifies the post-DCC observations. The baseline slope is *β*_1_, the post-intervention slope is *β*_1_ + *β*_3_, and the difference in slopes is *β*_3_.

The residual error component, *ε_t_*, was modelled using a SARIMA process, denoted SARIMA(*p*, *d*, *q*)(*P* , *D*, *Q*)*_s_*, to account for temporal autocorrelation and seasonal variation where appropriate. Differencing was incorporated where necessary to address non-stationarity. The adequacy of the temporal error structure was assessed using residual diagnostics. This is represented by the equation:

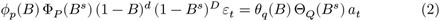

where *B* is the lag operator; *p*, *d*, *q* are the non-seasonal autoregressive, differencing and moving-average orders; *P* , *D*, *Q* are their seasonal equivalents; *s* is the seasonal period; and *a_t_* is a white-noise innovation.

The impact of the DCC is reported as two measures for each outcome. The first impact measure is on the *trend* difference: a comparison of the pre-DCC and post-implementation slopes to test whether implementation of the DCC had a statistically significant impact on the trend over time. The second impact measure is the *level* difference: the change in outcome from the end of the pre-DCC to the beginning of the post-implementation period, estimated from the linear model. Effect estimates were presented with 95% confidence intervals and p-values.

For hypothesis testing and 95% confidence intervals, following SARIMAX adjustment, a segmented joint linear regression model was fitted to the model-adjusted values to estimate the pre-DCC and post-implementation slopes, the difference between slopes, and the level change at the intervention boundary from the end of the pre-intervention to the start of the post-intervention periods. Heteroscedasticity-consistent (HC3) robust standard errors were calculated for the joint linear regression model. These robust standard errors were used to derive 95% confidence intervals and p-values for the estimated slopes, differences in slopes and level changes. Thus, SARIMAX modelling addressed the temporal dependence structure of the data, whereas HC3 robust standard errors were applied at the subsequent linear regression stage to account for potential heteroscedasticity. Model adequacy was evaluated using residual diagnostics, including tests of autocorrelation, normality and heteroscedasticity. All analyses were conducted in R version 4.4.2 (R Foundation for Statistical Computing, Vienna, Austria).

To interpret the magnitude of the intervention’s effect, we estimated the cumulative, annualised effect of the DCC during the observed post-implementation period. To do so, we imputed the model’s predicted average outcome per episode of care (e.g., bed-days per admission) for each week in the post-transition period with the DCC and without the DCC (the counterfactual projection assuming the pre-DCC trajectory had continued without the intervention) using the linear model fitted to the SARIMAX-adjusted values. Weekly differences were multiplied by the relevant weekly activity denominator (e.g., number of admissions), summed across the post-implementation period, annualised (e.g., bed-days per year), and expressed as both absolute units saved or gained, and relative percentage difference using the counterfactual as the denominator. The annualised figures thus represent the estimated, cumulative real-world difference integrated across all post-implementation weeks.

## Data Availability

The weekly averages metrics data that support the findings of this study are available in figshare at https://figshare.com/s/12ab5bafe561fb4c3ad1

## Code Availability

The underlying code for this study is available in figshare and can be accessed via this link https://figshare.com/s/12ab5bafe561fb4c3ad1.

## Acknowledgements

We would like to gratefully express our thanks to Kayley Lyons, who contributed to early design and conceptualisation of the evaluation, and Meredith Layton, who played a key convening and strategic role and offered valued comments on the manuscript.

The DCC implementation was funded by the RMH Foundation. This DCC evaluation study received no funding.

## Author contributions

BD performed data analyses, generated tables and figures, and wrote the initial manuscript draft. PYC provided supervision and coordination for the latter stages of the evaluation, provided methodological advice, reviewed statistical calculations, wrote sections of the manuscript and reviewed and edited multiple drafts. SP contributed to study conception, design and supervision, interpretation of findings, and critically reviewed and edited the manuscript. PS provided subject matter expertise and critically reviewed and edited the manuscript. SA contributed to the study conception, interpretation of findings, provided subject matter expertise, and critically reviewed the manuscript. BA and LS contributed to the study conception and design, interpretation of findings, and critically reviewed the manuscript. TS assisted with data analysis and critically reviewed the manuscript. TF provided the cleaned data, interpreted findings, contributed subject matter expertise and critically reviewed the manuscript. DC contributed to study design and conception, data analysis, interpretation of findings, and critically reviewed the manuscript. JG and WC contributed to study design and conception and critical review of the manuscript.

## Competing Interests

The authors declare no competing financial or non-financial interests.

Author Professor Peter Steel is Associate Editor of npj Health Systems. Professor

Steel was not involved in the journal’s review of, or decisions related to, this manuscript.

