## Supplementary material for "Impact of a hospital digital coordination centre on patient flow in a large tertiary centre"

### Supplementary materials

#### S1: Illustrating low values at the start of the pre-DCC period for escalation metrics

*Figure S1. Change in weekly percent of 15-minute blocks where the Ambulance Victoria and waiting room queues comprising 6 to 19 patients, before and after DCC implementation, and expected values had the DCC not been implemented (the counterfactual trend)*

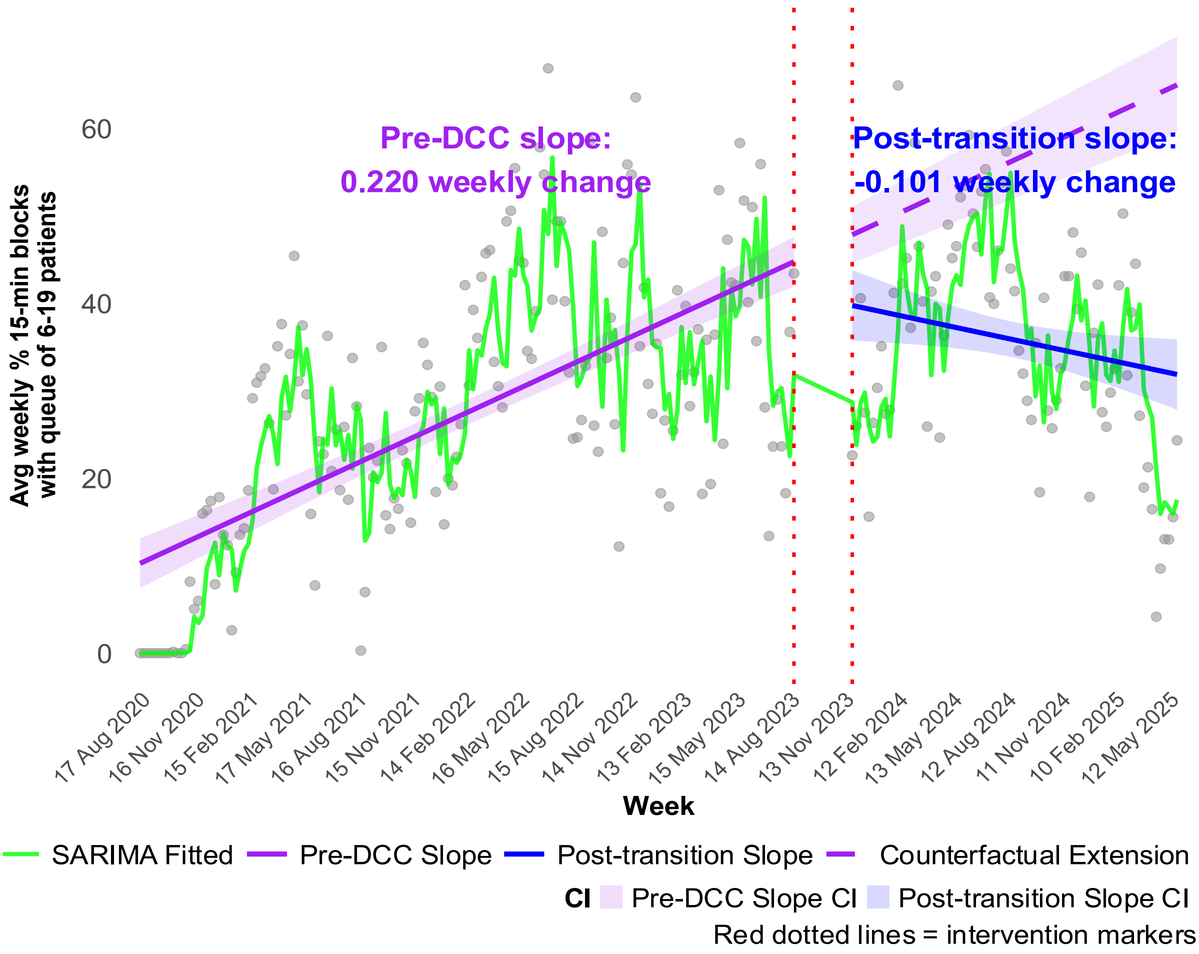

*Figure S2. Change in the weekly mean percentage of 15-minute blocks where bed variance was 10 to 19 patients, before and after DCC implementation, and expected values had the DCC not been implemented (the counterfactual trend)*

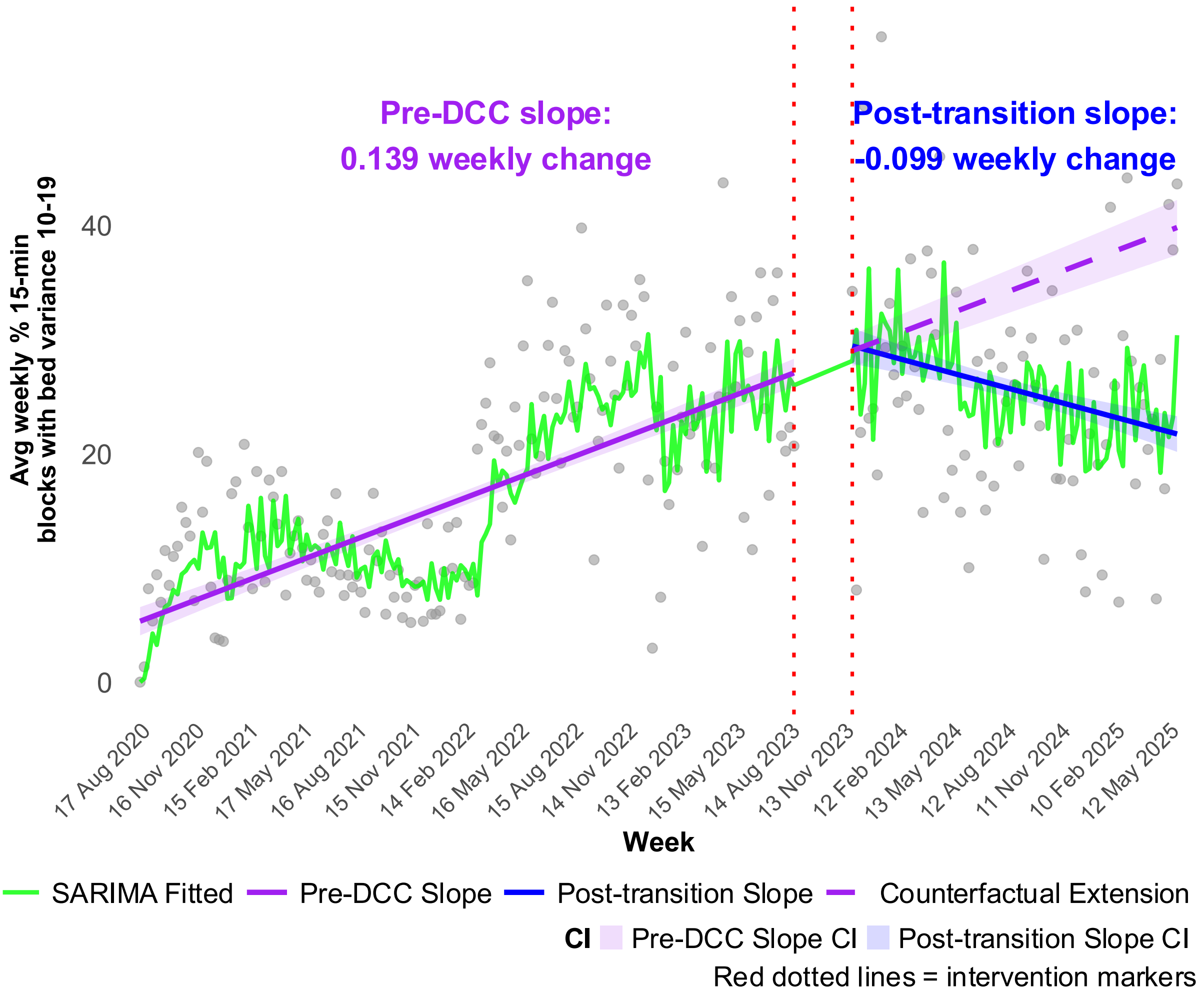

#### S2. Escalation triggers

The following table indicates the agreed metrics which form triggers and thresholds used to identify excessive demand. Further details can be found at RMH Capacity and Patient Flow Escalation Procedure (Royal Melbourne Hospital, n.d.).

*Table S1: Capacity management levels and escalation triggers at Royal Melbourne Hospital*

| Level 0 Capacity Management triggers | Level 1 escalation triggers * | | Level 2 escalation triggers * |
|---|---|---|---|
| Bed variance 0-9 | Bed variance 10-19 | | Bed variance ≥ 20 |
| ≤ 5 Beds closed | 6-9 Beds closed | | ≥10 Beds closed |
| ≤ 65 patients in ED | 66- 94 patients in ED | | ≥ 95 patients in ED |
| ≤ 20 patients in ED awaiting an inpatient bed | 21-35 patients in ED awaiting an inpatient bed | | ≥ 36 patients in ED awaiting an inpatient bed |
| 0- 5 AV /WRQ | 6- 19 in AV/WRQ | | ≥ 20 patients in AV/WRQ |
| **Level 0 Capacity Management** | **Level 1 escalation** | | **Level 2 escalation** |

#### S3: Average weekly ED arrivals and inpatient admissions for all available data

*Figure S3. Time-series plot of average weekly ED arrivals from 2020 to 2026 (red line indicates where data analysis began)*

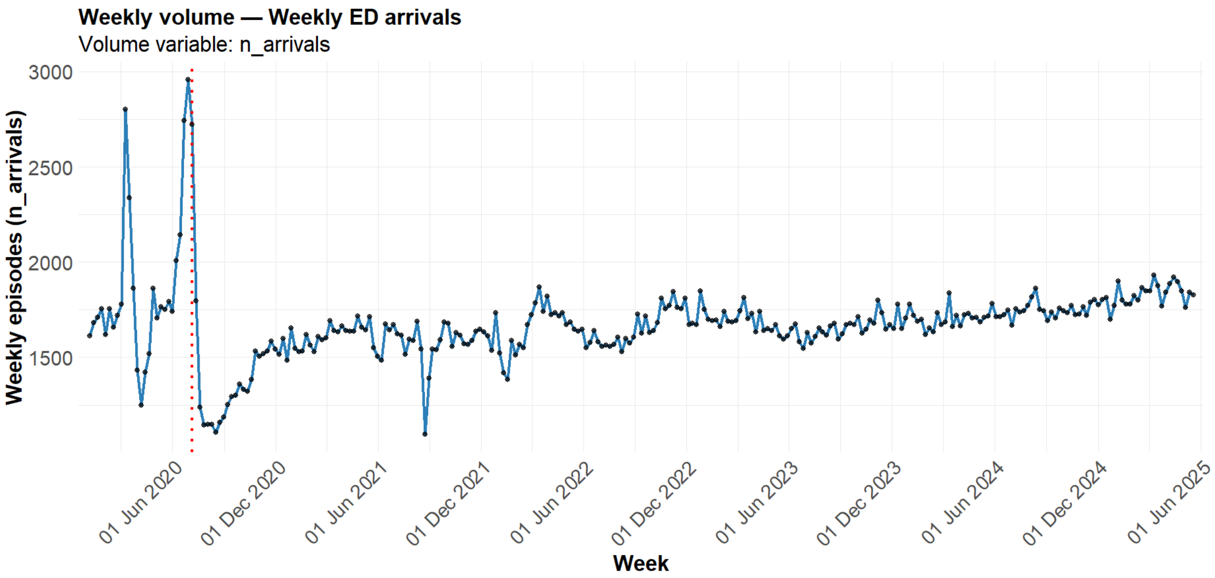

*Figure S4. Time-series plot of average weekly inpatient admissions from 2020 to 2026 (red line indicates where data analysis began)*

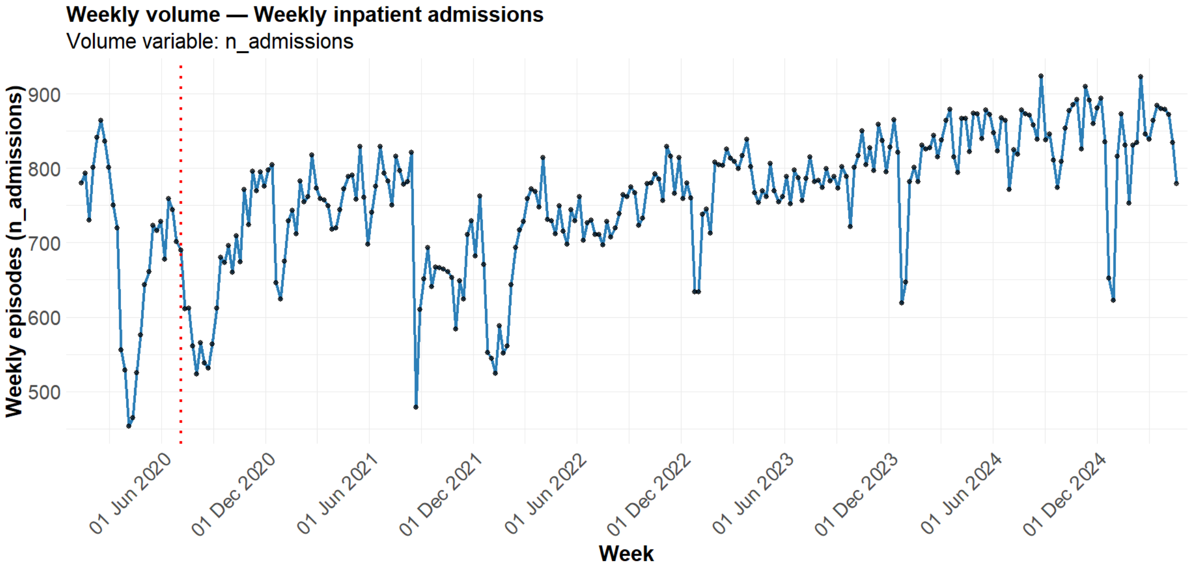

#### S4: Model diagnostics and assumption testing

Diagnostic testing indicated that the SARIMAX models adequately captured the temporal structure of the weekly patient flow metrics. Across all analysed outcomes—including inpatient length of stay, emergency department length of stay, percentage of discharges prior to 12 pm, proportion admitted within four hours, and escalation trigger metrics—residual time plots showed no systematic temporal trends following model fitting. Autocorrelation and partial autocorrelation functions of the residuals demonstrated that remaining correlations were small and fell within expected bounds, suggesting that the autoregressive and moving-average components sufficiently addressed serial dependence in the data.

Stationarity assumptions were satisfied following the differencing structure applied to each time series. Residual distributions were centred around zero with broadly stable variance across the study period. Minor deviations from normality and mild heteroscedasticity were observed in some models, which is common in healthcare operational data; therefore, robust standard errors were used when estimating model coefficients and confidence intervals. Multicollinearity was over 0.8 for some analyses, but repeating the analyses after centring time resulted in very similar results, so centering was avoided to allow ease of interpretation. Overall, these diagnostic results indicate that the model assumptions were adequately met and support the validity of the estimated level and slope changes associated with implementation of the Digital Coordination Centre.

Table S2. summarises model selection and assumption testing diagnostics. Detailed diagnostic plots, including residual time series and residual distribution checks for each outcome metric, are provided in the Supplementary Materials.

*Table S2. Final SARIMAX model specifications and key diagnostics for each analysed metric*

| Outcome metric | SARIMAX (p,d,q) (P,D,Q)[s] | Seasonality selected | Final AIC | Ljung-Box p | Anderson-Darling normality test | Breusch-Pagan p | Overall validity |
|---|---|---|---|---|---|---|---|
| **ED LoS** | (3,1,1) (0,0,1)[*13*] | Quarterly (13 weeks) | 436.4 | 0.54 | 0.06 | <0.01 | Valid using robust SEs |
| **Admitted within 4 hours (NEAT)** | (0,1,2) (1,0,0)[*52*] | Yearly (52 weeks) | 1423.56 | 0.70 | 0.79 | 0.19 | Fully valid |
| **Inpatient LoS** | (0,1,1) (1,0,0)[*52*] | Yearly (52 weeks) | -27.26 | 0.88 | 0.02 | 0.37 | Valid using robust SEs |
| **Discharged before 12 pm** | (1,1,1) (1,0,1)[*13*] | Quarterly (13 weeks) | 1055.50 | 0.54 | 0.01 | 0.05 | Valid using robust SEs |
| **% of 15-min blocks with Ambulance Victoria / wait room queue of 6–19** | (1,1,1) (0,0,1)[*52*] | Yearly (52 weeks) | 1758.84 | 0.15 | 0.62 | <0.01 | Valid using robust SEs |
| **% of 15-min blocks with Ambulance Victoria / wait room queue of ≥20** | (0,1,1) (0,0,0) | Non-seasonal | 1066.52 | 0.54 | <0.01 | <0.01 | Valid using robust SEs |
| **% of 15-min blocks with bed variance 10–19** | (0,1,5) (0,0,0) | Non-seasonal | 1654.50 | 0.57 | <0.01 | <0.01 | Valid using robust SEs |
| **% of 15-min blocks with bed variance ≥20** | (2,1,1) | Non-seasonal | 1803.39 | 0.29 | 0.01 | <0.01 | Valid using robust SEs |

**All models were estimated using SARIMAX with an exogenous intervention term representing the pre-DCC and post-implementation periods. For non-seasonal models, seasonal parameters were set to zero. Ljung-Box tests assessed residual autocorrelation; Anderson-Darling assessed residual normality; and Breusch-Pagan and White tests assessed heteroscedasticity. Because several models showed evidence of non-normality and/or heteroscedasticity, robust HC3 standard errors were used for inference.**

LoS = length of stay

SARIMAX = seasonal autoregressive integrated moving average time-series analyses with an exogenous variable

#### Residual and auto-correlation plots for each outcome

*Figure S5: Diagnostic plots for SARIMAX analysis of average weekly percent of 15-min blocks where Ambulance Victoria / wait room queue is 6-19*

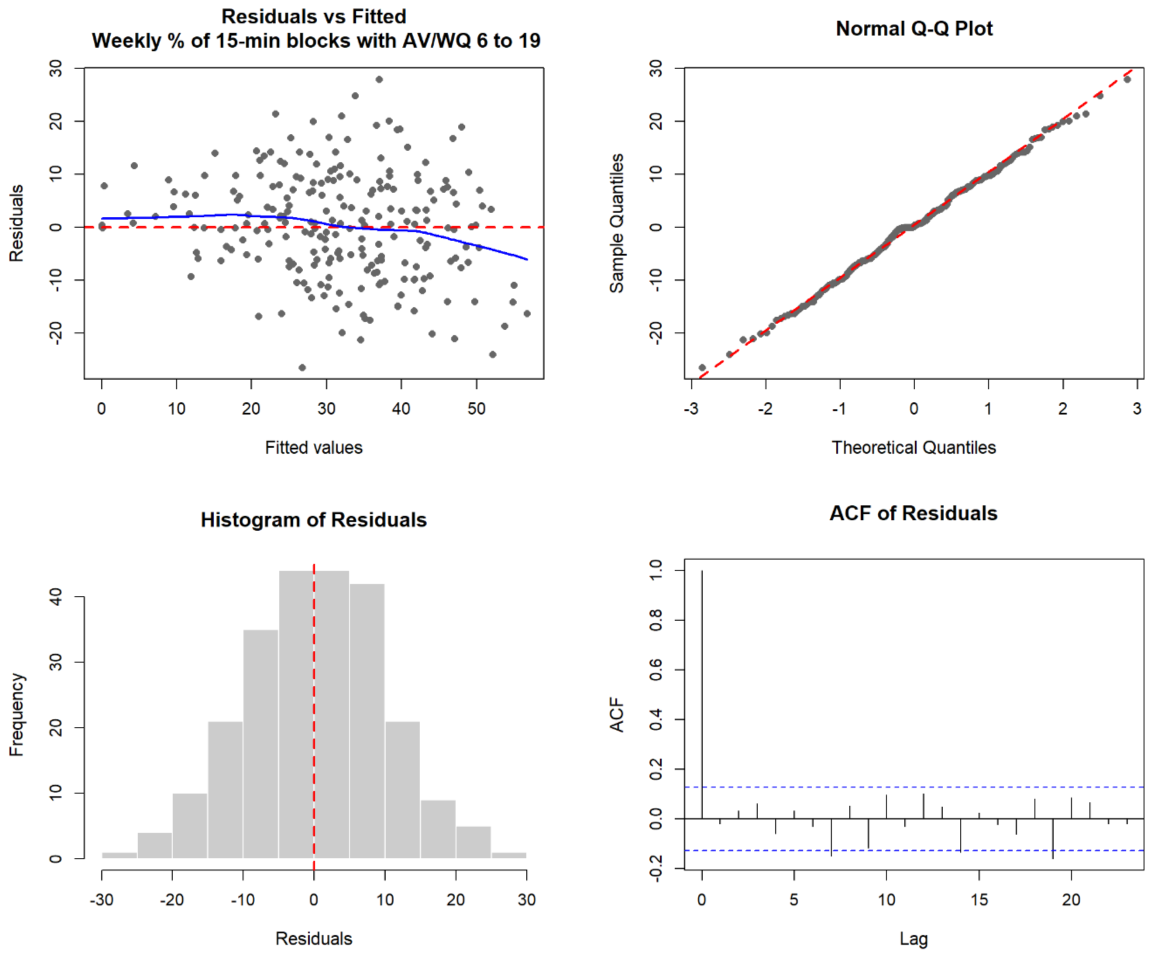

*Figure S6: Diagnostic plots for SARIMAX analysis of average weekly percent of 15-min blocks were Ambulance Victoria / wait room queue is 20 or greater*

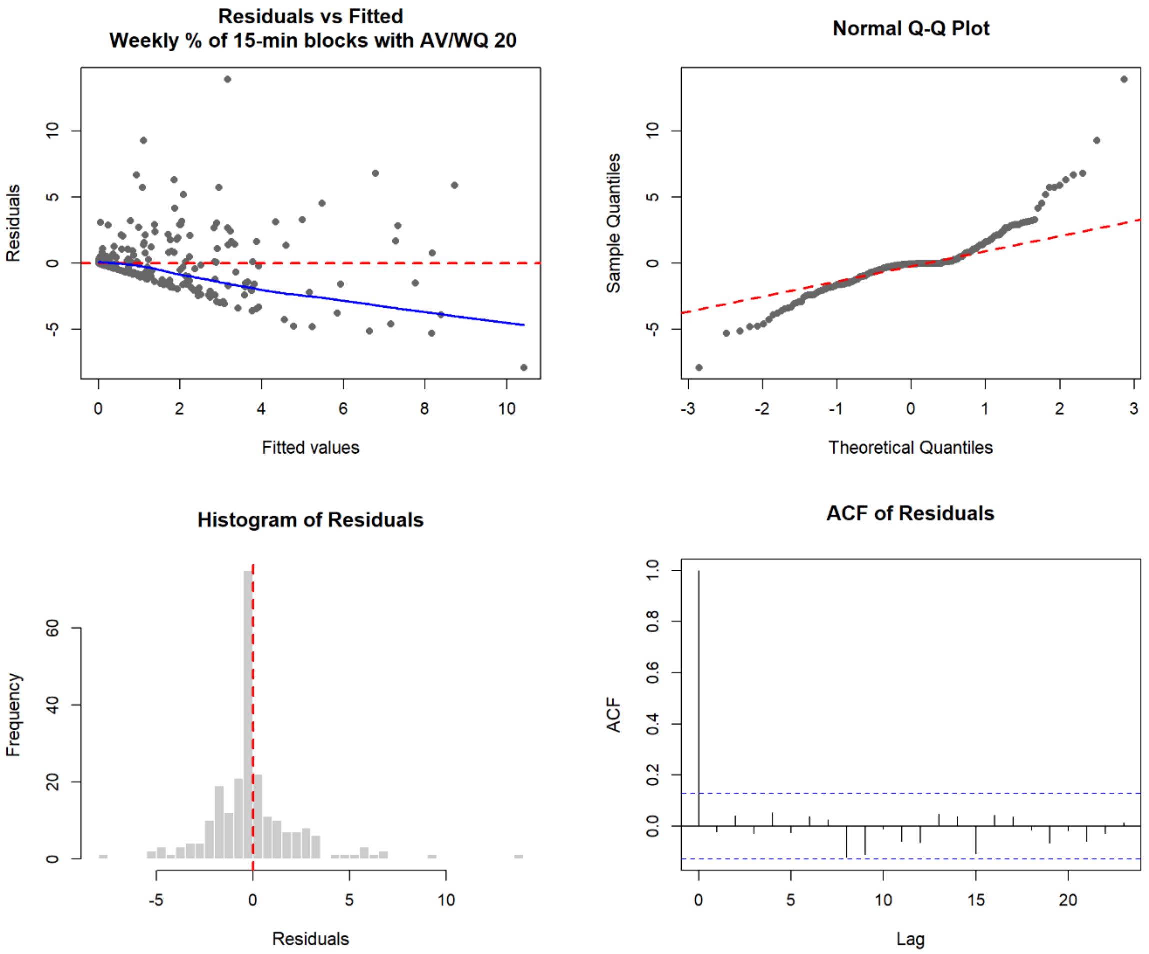

*Figure S7: Diagnostic plots for SARIMAX analysis of average weekly percent of 15-min blocks where bed variance is 10-19*

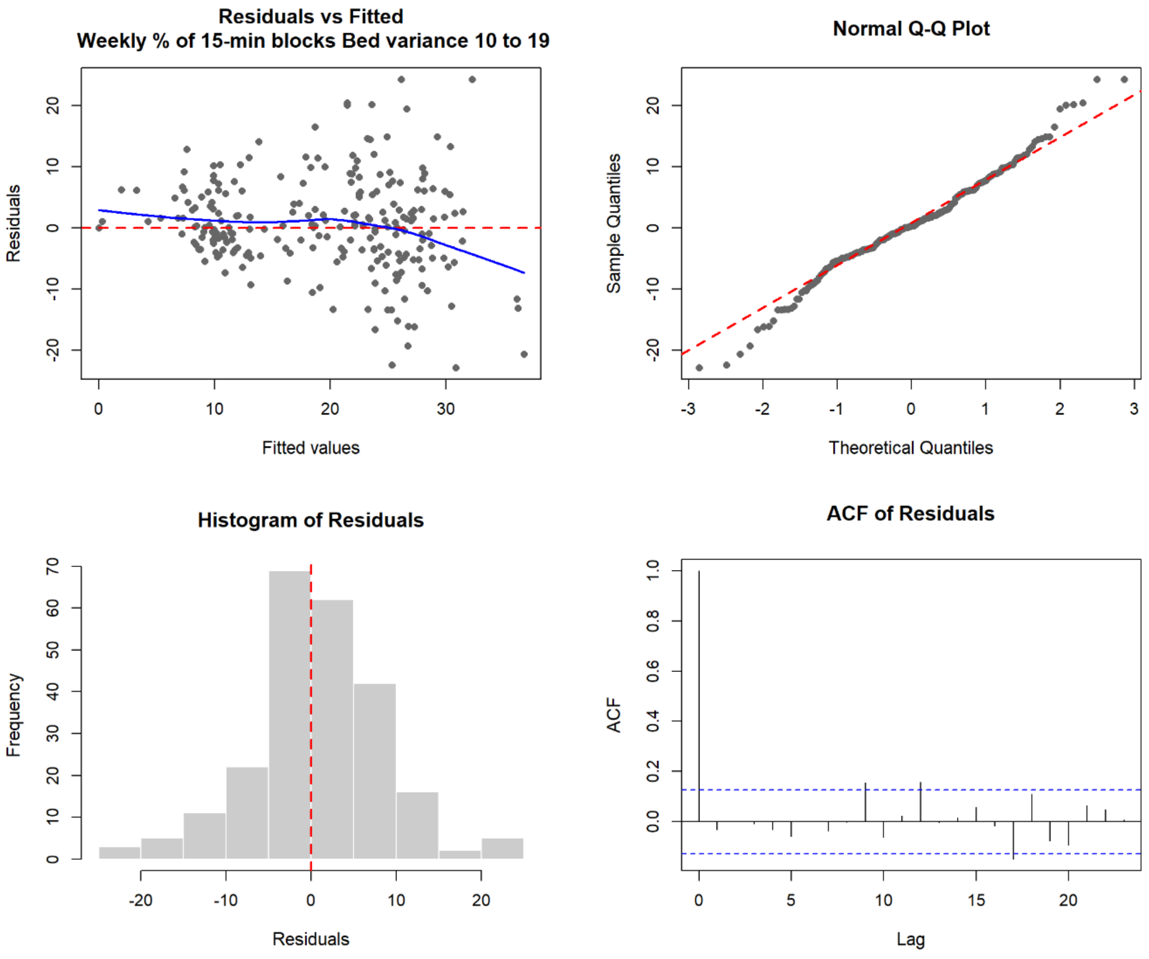

*Figure S8: Diagnostic plots for SARIMAX analysis of average weekly percent of 15-min blocks where bed variance is 20 or greater*

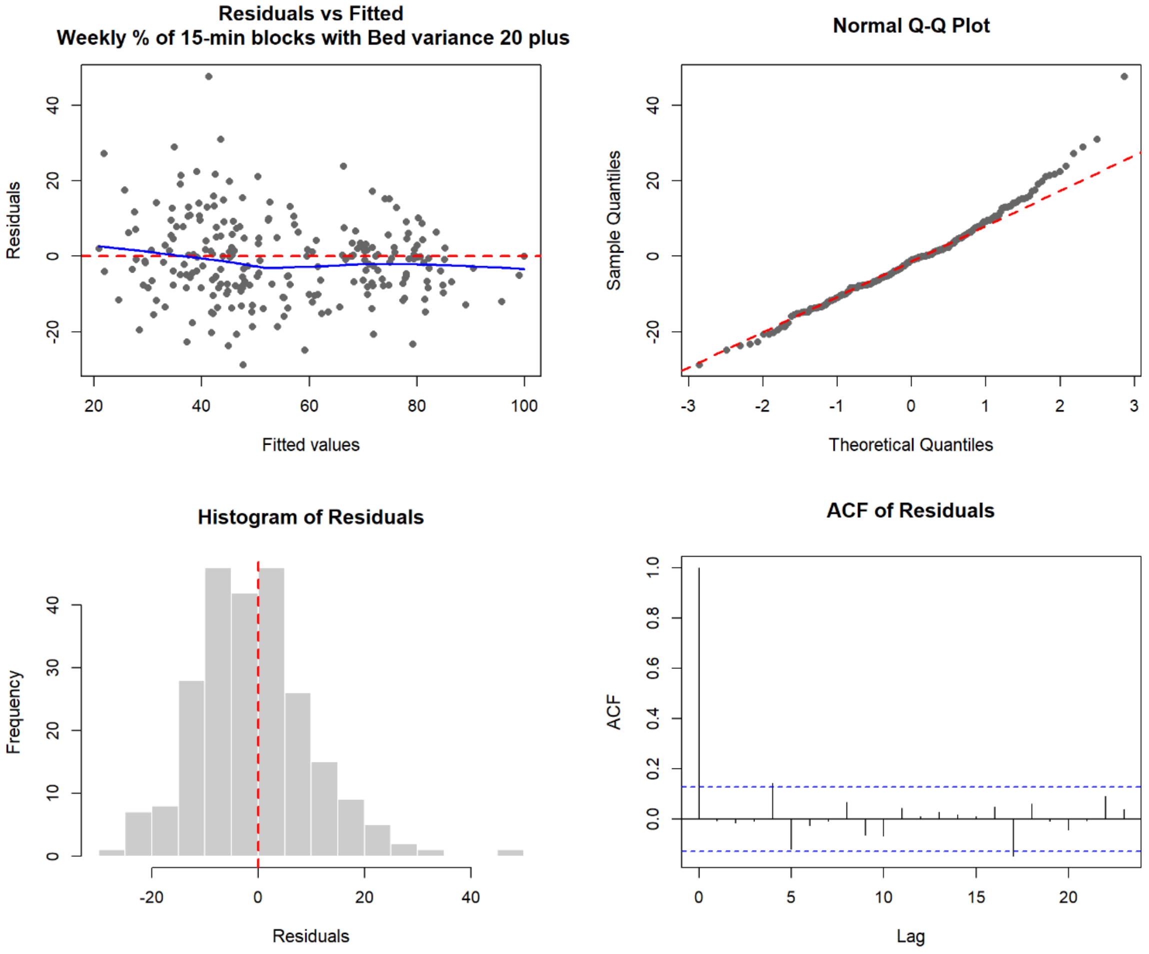

*Figure S9: Diagnostic plots for SARIMAX analysis of average weekly emergency department length of stay*

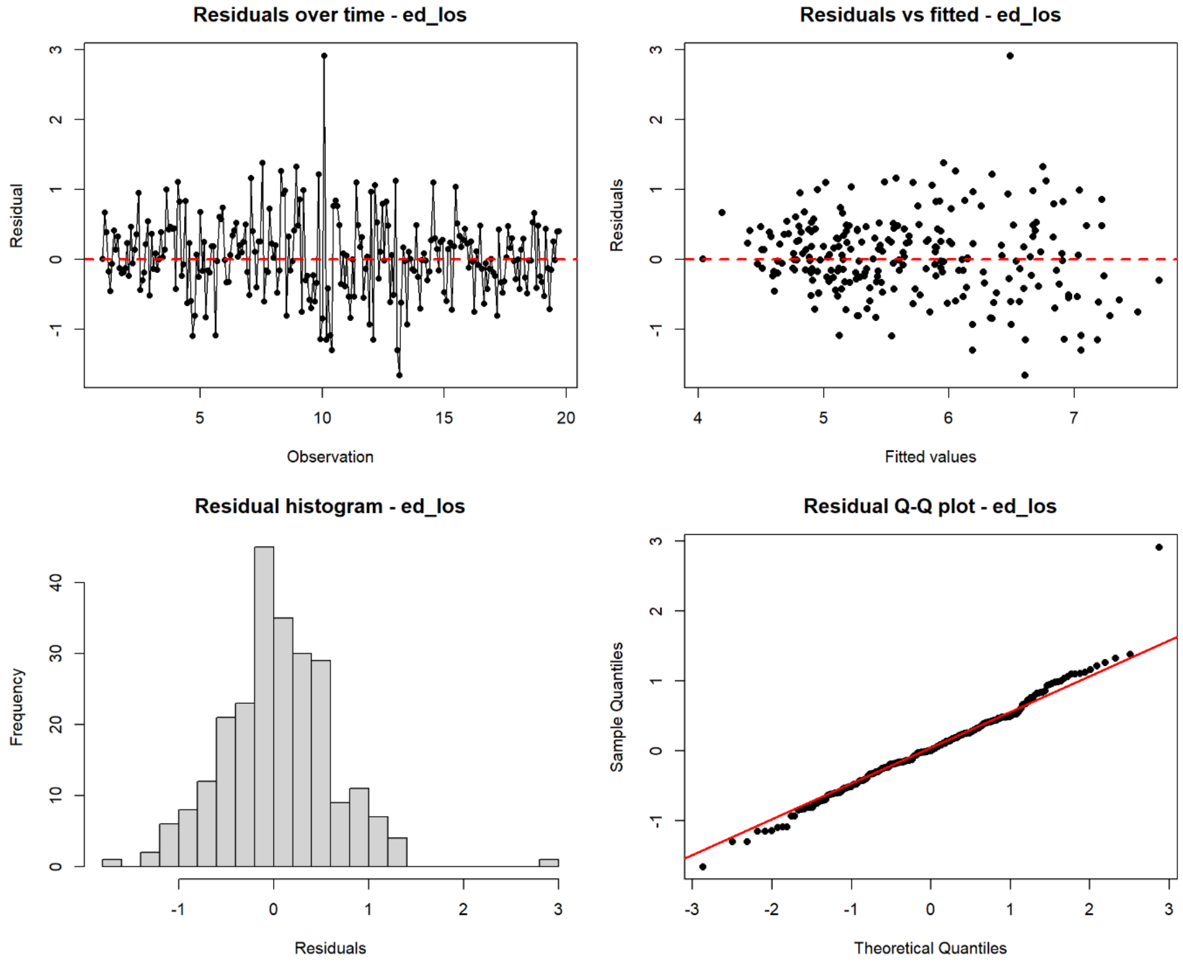

*Figure S10: Diagnostic plots for SARIMAX analysis of average weekly proportion of admissions with 4 hours (NEAT)*

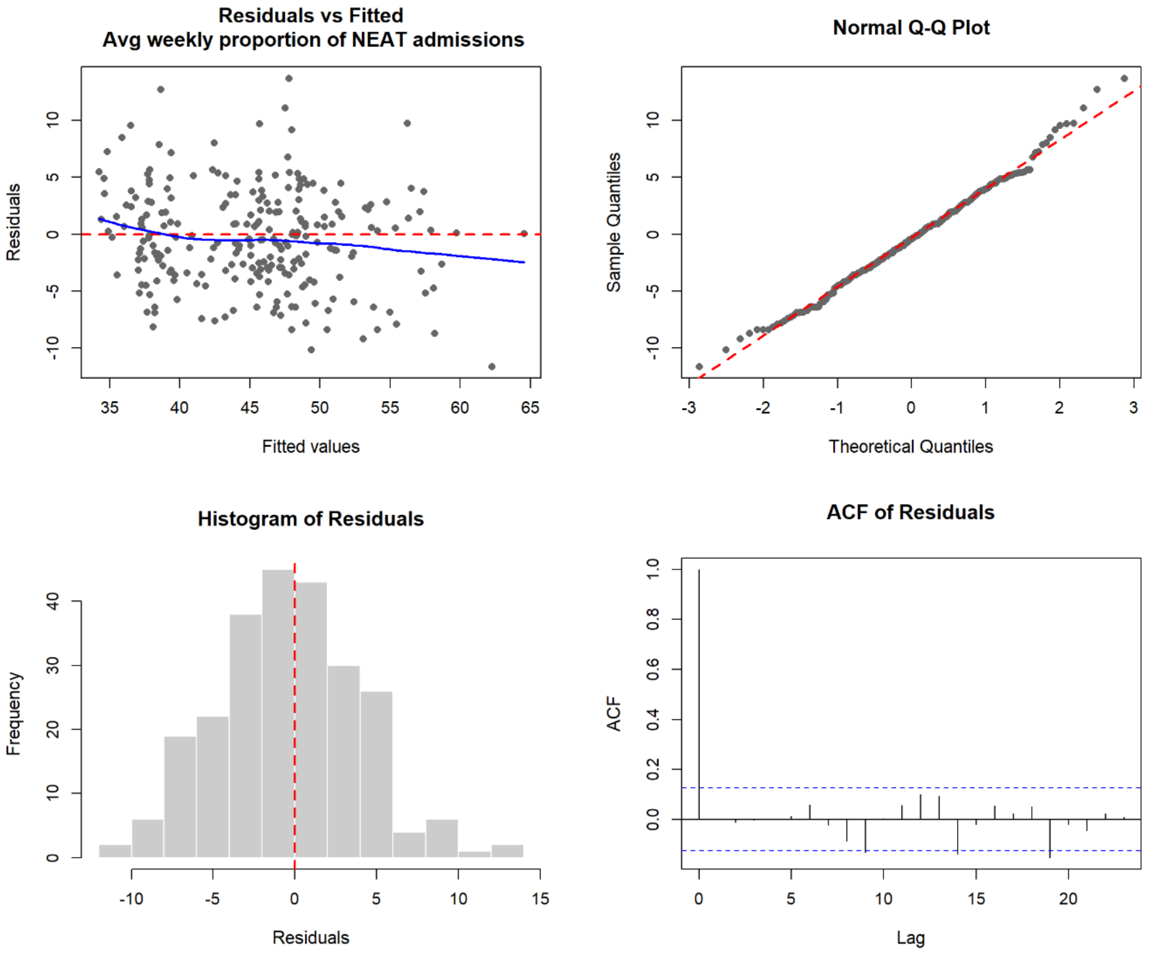

*Figure S11: Diagnostic plots for average weekly for SARIMAX analysis of inpatient length of stay*

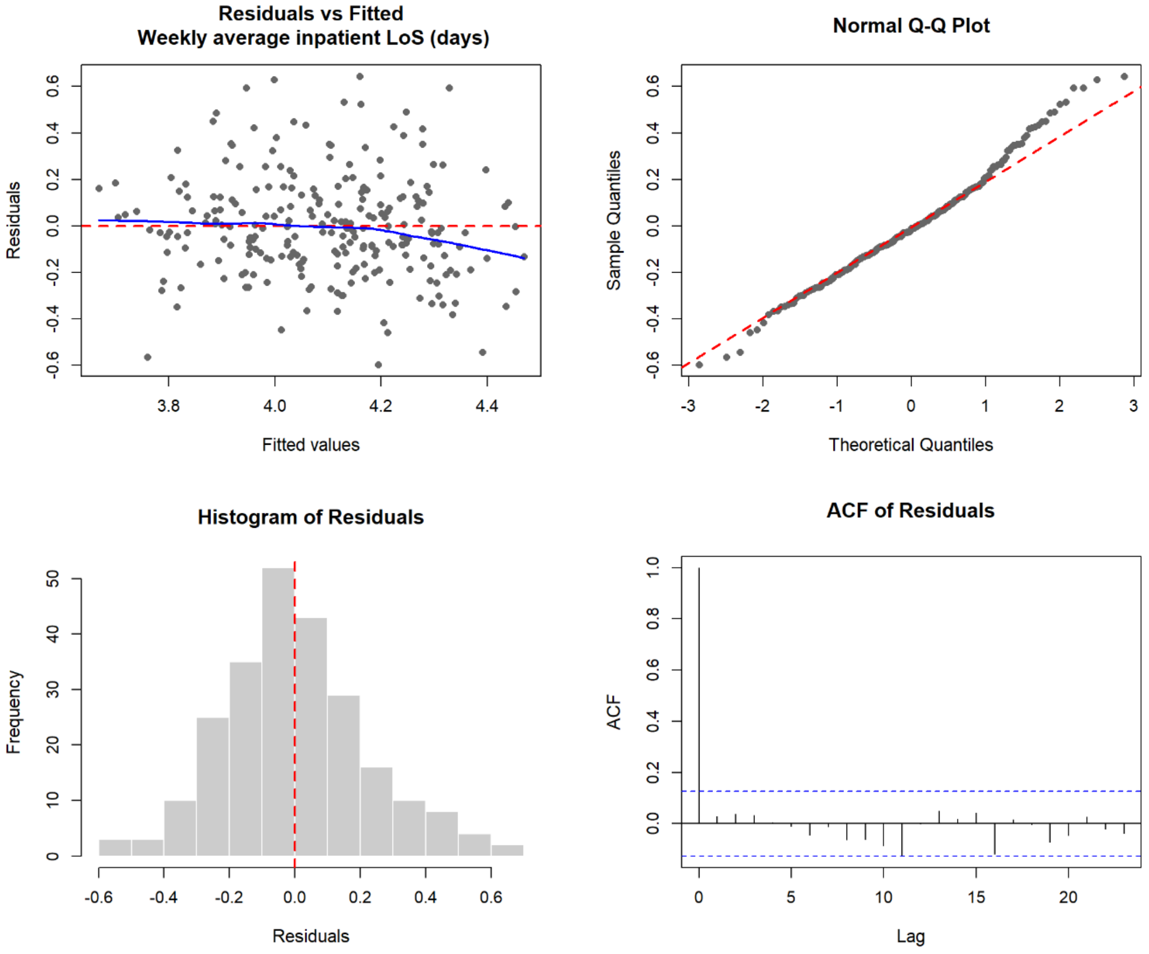

*Figure S12: Diagnostic plots for SARIMAX analysis of average weekly proportion of discharges by 12pm*

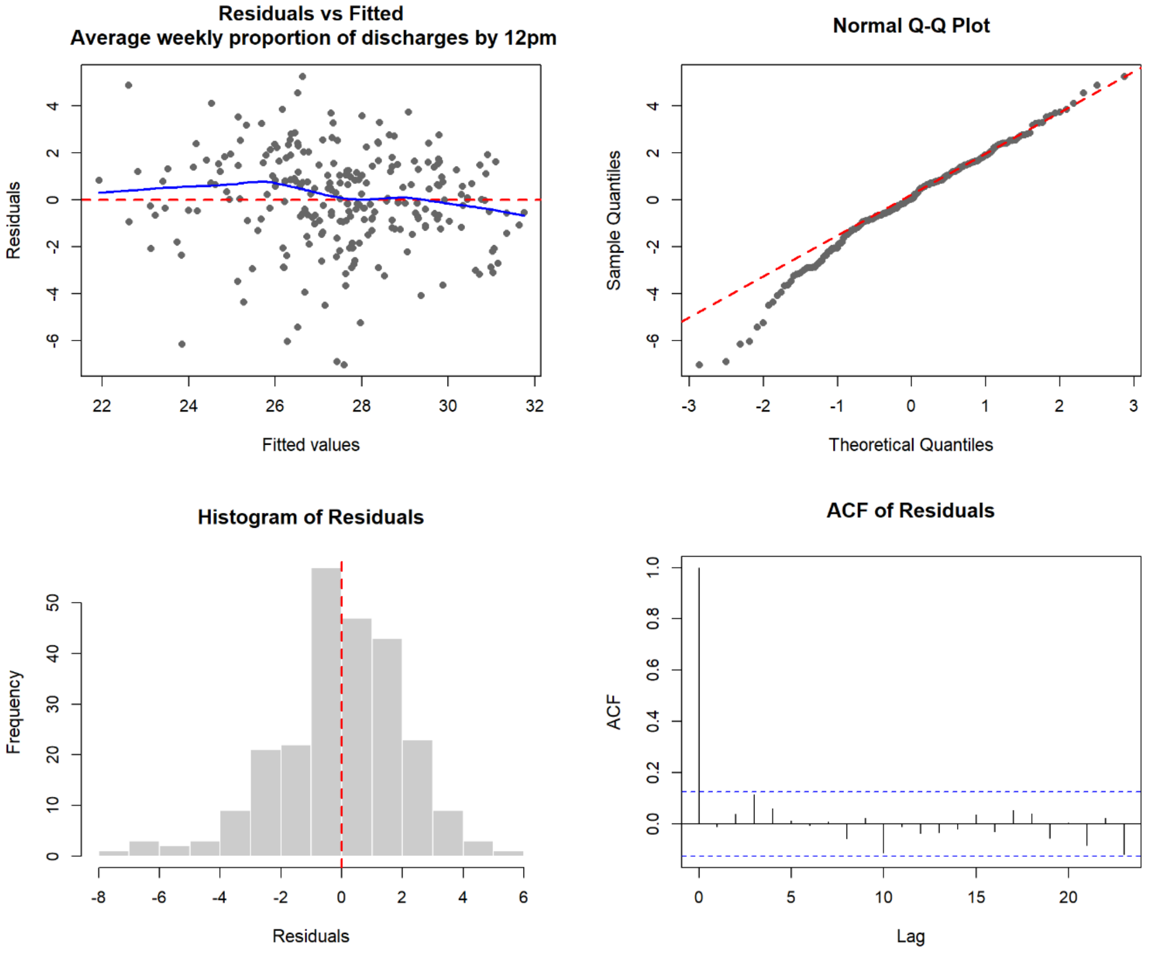

#### S4: Utilising an earlier start date for the omitted DCC implementation period

Table S3 provides the results of a sensitivity analysis whereby the omitted DCC implementation period was changed to begin on February 13 2023, rather than November 28$^{th}$ 2023, in order to omit all periods where interventions were implemented eg when a Patient Flow Co-ordinator team was introduced, a purpose-built transit lounge was opened, and digital workflows and real-time dashboards were implemented. Results were similar to the original analyses except they were a little larger overall.

*Table S3: Results of sensitivity analysis where the date for the DCC implementation period omitted from analyses was changed to Feb 13, 2023*

| | **Intervention effect** | | | | | | | | | |
|---|---|---|---|---|---|---|---|---|---|---|
| | **Original analysis: Post-DCC starts at Nov 27th, 2023** | | **Sensitivity analysis: Post-DCC starts at Feb 13th, 2023** | | **Original analysis: Post-DCC starts at Nov 27th, 2023** | | **Sensitivity analysis: Post-DCC starts at Feb 13th, 2023** | | **Original counterfactual different** | **Sensitivity analysis: Counterfactual different** |
| | **Difference in slopes (CI)** | **$Pr\|H_0$** | **Difference in slopes (CI)** | **$Pr\|H_0$** | **Level change (CI)** | **$Pr\|H_0$** | **Level change (CI)** | **$Pr\|H_0$** | **% changed (annualised difference)** | |
| **Escalation metrics** | | | | | | | | | | |
| Ambulance Vic & waiting room queue of 6–19 | −0.27 (−0.37, −0.17) | <.001 | −0.19 (−0.28, −0.09) | <.001 | −5.07 (−10.50, 0.35) | 0.067 | −15.88 (−22.22, −9.54) | <.001 | 30% decrease (~5,400 fewer triggers/year) | 37% decrease (~7,700 weighted units/year) |
| Ambulance Vic & waiting room queue of 20+ | −0.01 (−0.03, 0.01) | .499 | 0.004 (−0.02, 0.03) | .691 | 0.68 (−0.46, 1.82) | 0.242 | −1.05 (−2.16, 0.06) | .062 | NS | NS |
| Bed variance 10-19 | −0.23 (−0.27, −0.19) | <.001 | −0.23 (−0.28, −0.18) | <.001 | 1.07 (−1.05, 3.19) | 0.320 | −0.96 (−3.89, 1.961) | .517 | 24% decrease (~2,700 fewer triggers/year) | 26% decrease (~3,100 weighted units/year) |
| Bed variance 20+ | 0.03 (−0.02, 0.09) | .239 | 0.04 (−0.04, 0.11) | .337 | 6.23 (3.06, 9.41) | <0.001 | 5.07 (0.16, 9.98) | .043 | 25% increase (~2,600 additional triggers/year) | 19% increase (~2,200 weighted units/year) |
| **Emergency department metrics** | | | | | | | | | | |
| Weekly mean ED length of stay (hours) | −0.02 (−0.02, −0.015) | <0.001 | −0.02 (−0.03, −0.02) | <0.001 | −1.84 (−2.10, −1.58) | <0.001 | −2.30 (−2.60, −1.99) | <0.001 | 33% decrease (~119,000 fewer hours/year) | 38% decrease (~146,000 weighted hours/year) |
| Weekly % admitted within 4 hours | 0.12 (0.10, 0.14) | <0.001 | 0.15 (0.13, 0.17) | <0.001 | 15.02 (13.52, 16.52) | <0.001 | 18.20 (16.47, 19.93) | <0.001 | 73% increase (~9,100 additional timely admissions/year) | 105% increase (~11,000 weighted units/year) |

| | | | | | | | | | | |
|---|---|---|---|---|---|---|---|---|---|---|
| **Inpatient metrics** | | | | | | | | | | |
| Weekly mean inpatient length of stay (days) | −0.004 (−0.006, −0.003) | <0.001 | −0.004 (−0.005, −0.003) | <.001 | −0.16 (−0.22, −0.09) | <0.001 | −0.26 (−0.32, −0.19) | <0.001 | 8% decrease (~13,900 fewer inpatient days/year) | 9% decrease (~17,300 weighted days/year) |
| Weekly % discharged by 12pm | 0.01 (0.00, 0.02) | 0.018 | 0.01 (−0.005, 0.02) | .267 | 0.90 (0.28, 1.51) | 0.004 | 1.14 (0.27, 2.01) | .011 | 5% increase (~700 additional early discharges/year) | 5% increase (~690 weighted units/year) |

Notes: ED – emergency department; Post-DCC – post-implementation period; CI – 95% confidence interval; $Pr|H_0$ – probability of the observed or more extreme results, given that the null hypothesis is true.

*Figure 13. Change in weekly percent of 15-minute blocks where the Ambulance Victoria and waiting room queues comprising 6 to 19 patients, before and after DCC implementation, and expected values had the DCC not been implemented (the counterfactual trend)*

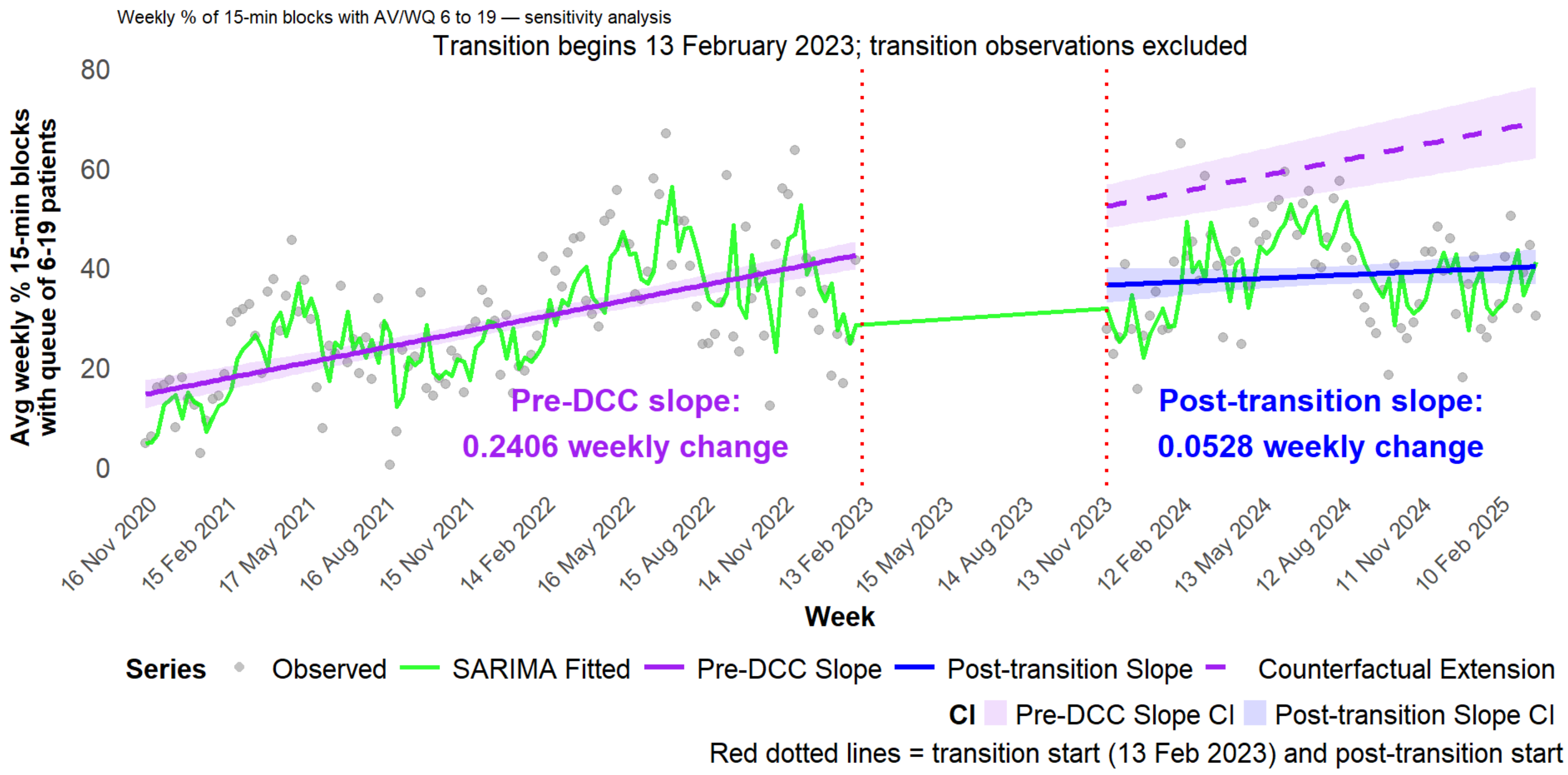

*Figure S14. Change in weekly percentage for 15-minute blocks where the Ambulance Victoria and waiting room queues comprised 20 patients or more, before and after DCC implementation, and expected values had the DCC not been implemented (the counterfactual trend)*

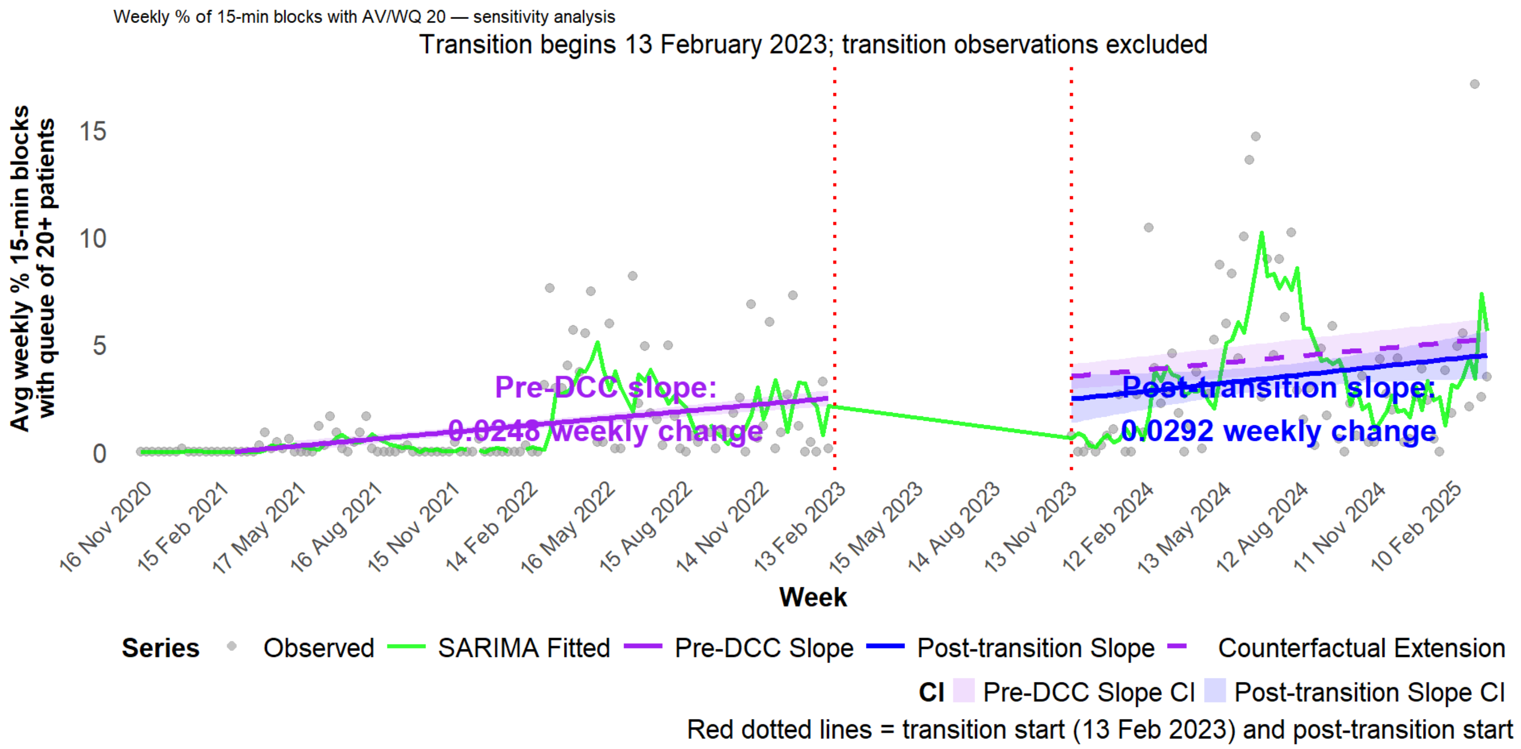

*Figure S15. Change in the weekly mean percentage of 15-minute blocks where bed variance was 10 to 19 patients, before and after DCC implementation, and expected values had the DCC not been implemented (the counterfactual trend)*

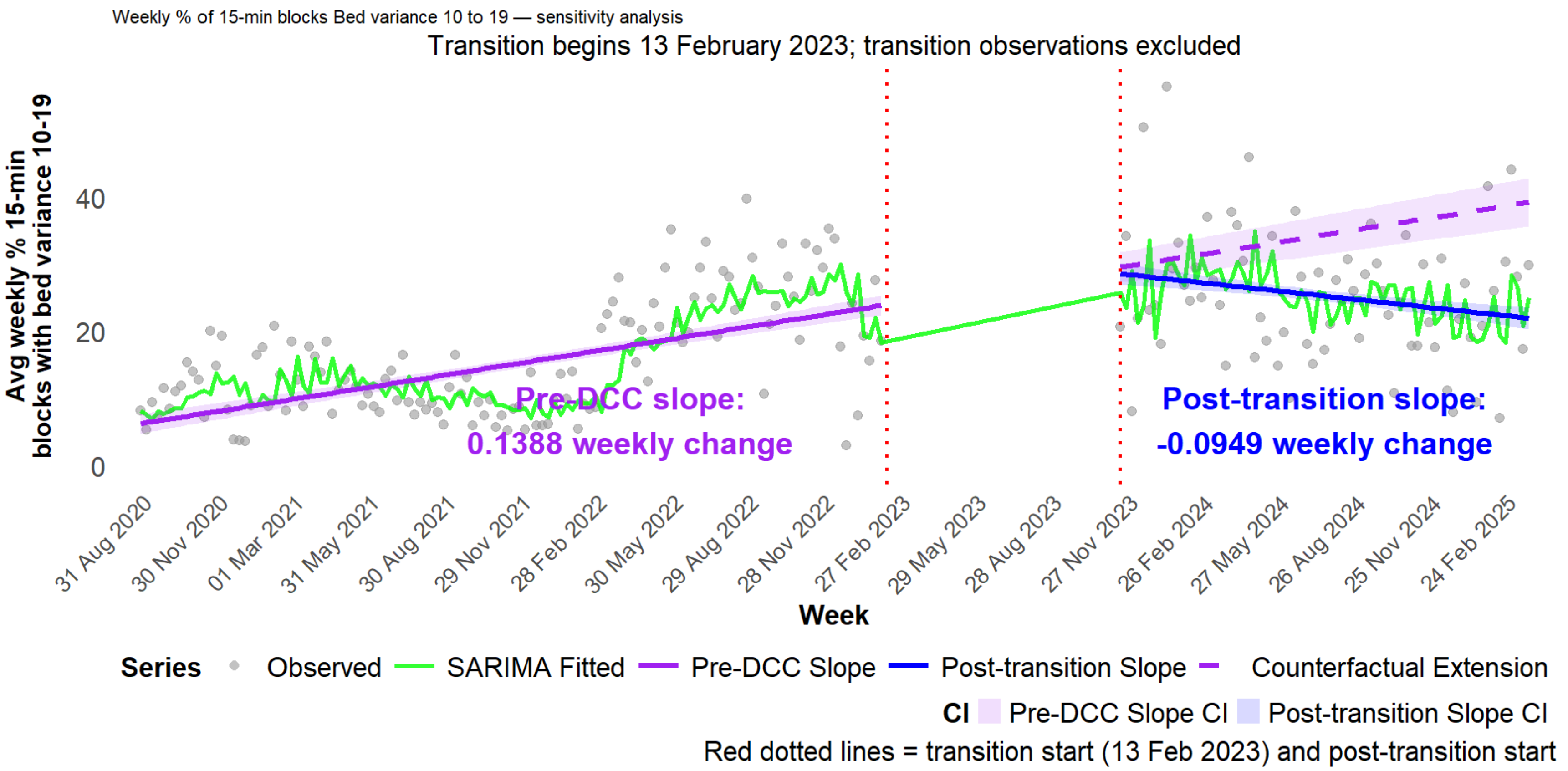

*Figure S16. Change in weekly percentage for 15-minute blocks where bed variance was 20 or greater, before and after DCC implementation, and expected values had the DCC not been implemented (the counterfactual trend)*

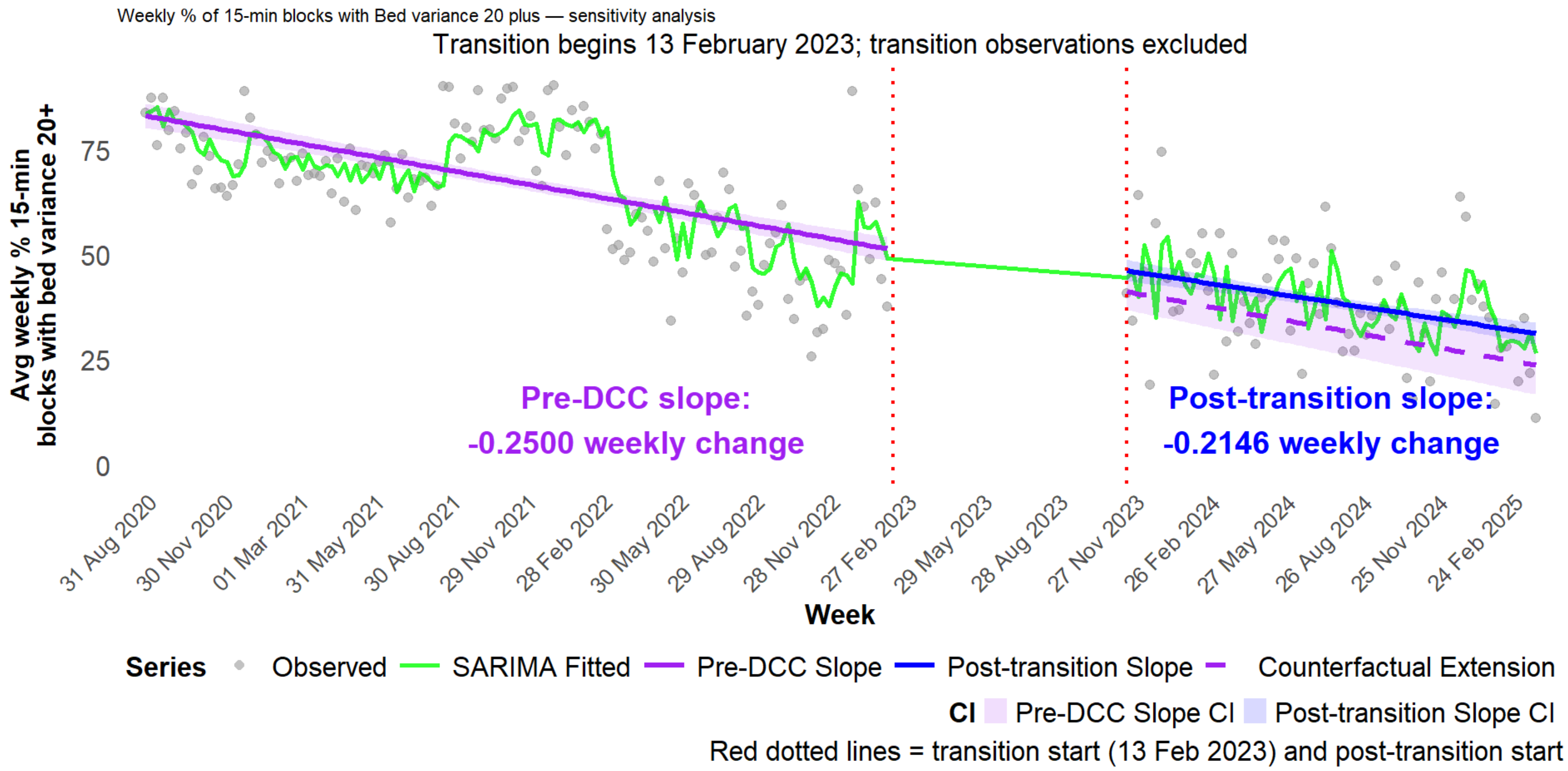

*Figure S17. Change in weekly average emergency department length of stay per week, before and after DCC implementation, and expected values had the DCC not been implemented (the counterfactual trend)*

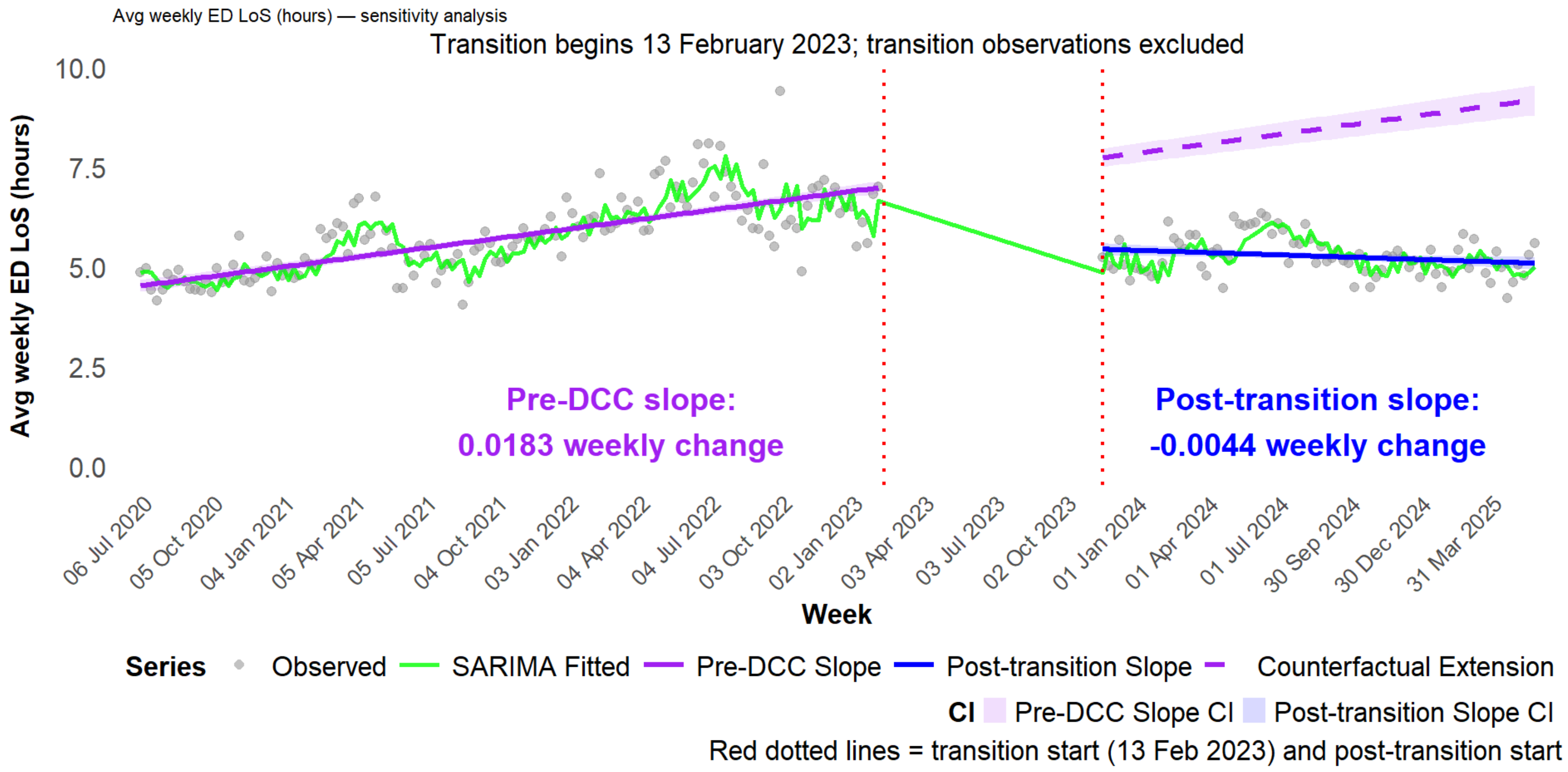

*Figure S18. Change in weekly percentage of ED patients admitted within 4 hours of presentation, before and after DCC implementation, and expected values had the DCC not been implemented (the counterfactual trend)*

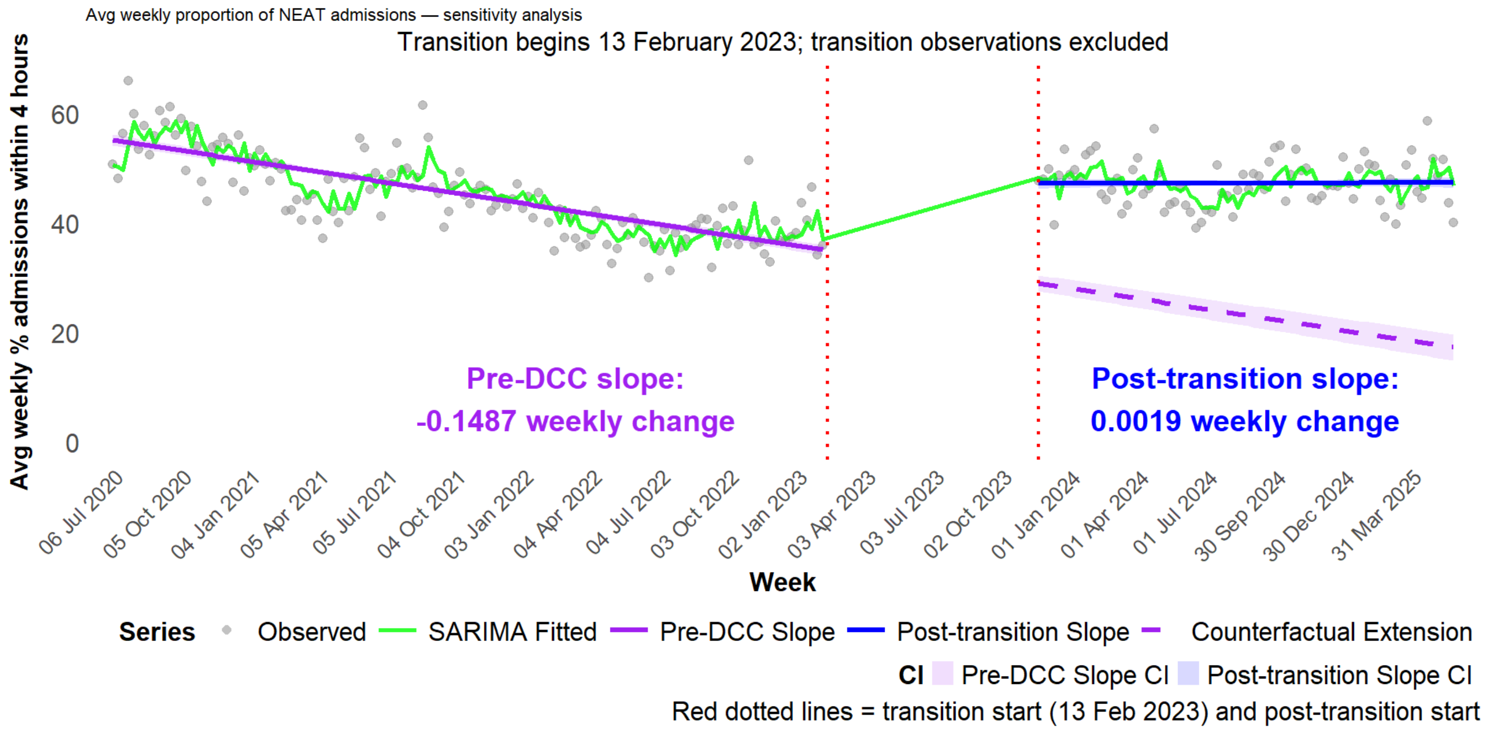

*Figure S19. Change over time in average weekly inpatient length of stay before and after DCC implementation, and expected values had the DCC not been implemented (counterfactual trend)*

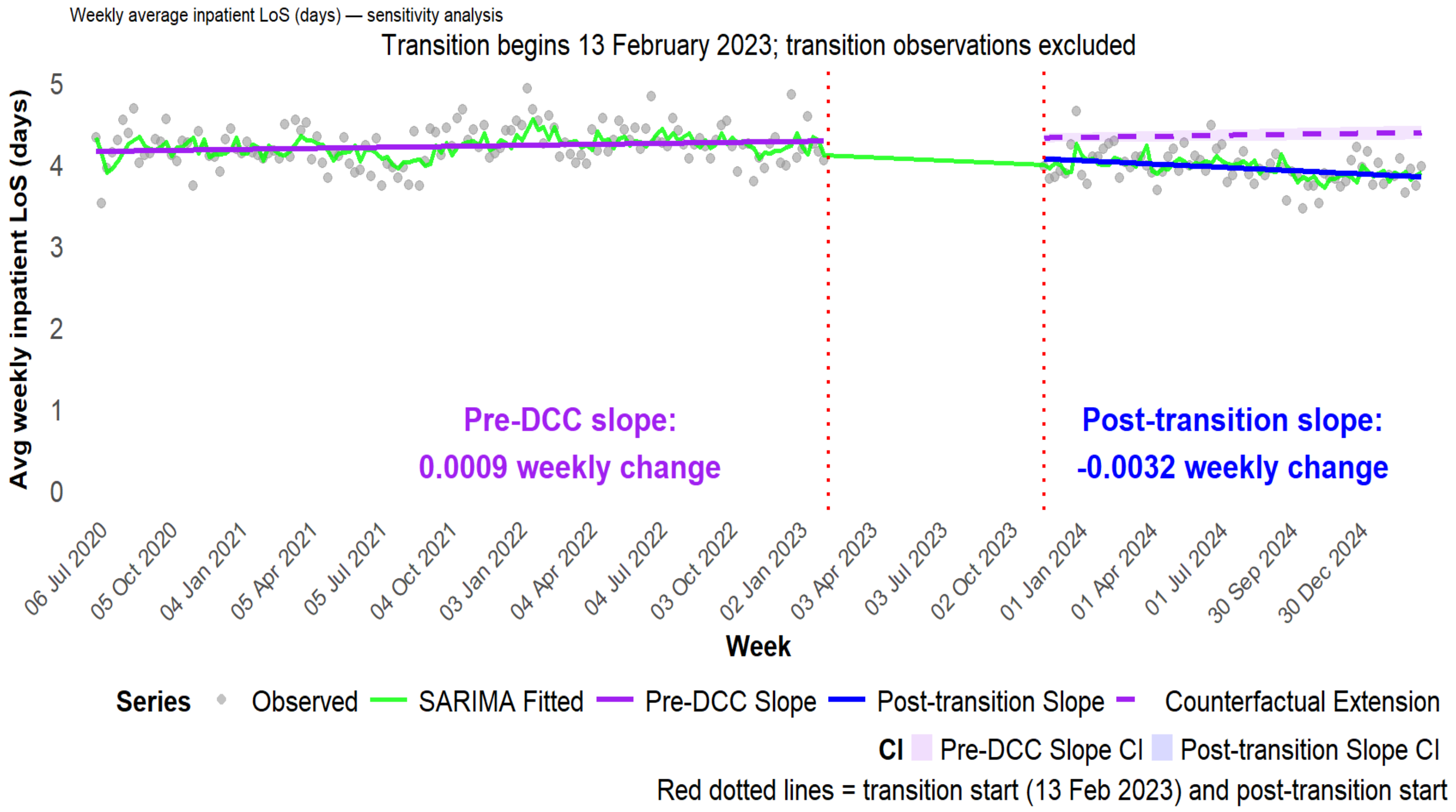

*Figure S20. Weekly percent of discharges prior to 12pm before and after DCC implementation and imputed values had the DCC not been implemented*

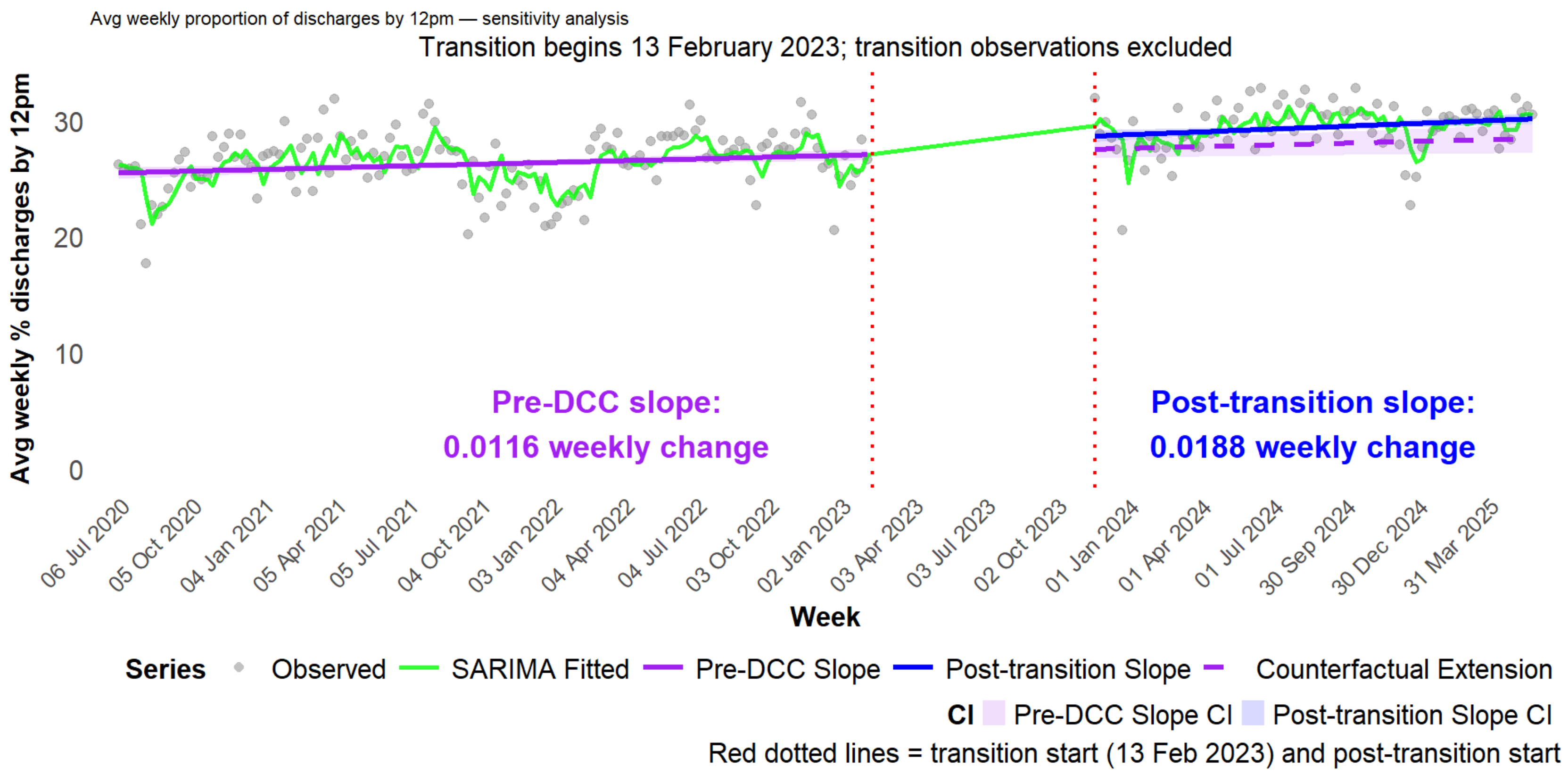
